# Anillin variant in proteinuric kidney disease drives tubular epithelial cell death, junctional instability, and barrier dysfunction

**DOI:** 10.64898/2026.03.24.26348901

**Authors:** Zie Craig, Holly M. Jacobs, Damian Fermin, Matthew Fischer, Xiaoyun Liu, Celine C. Berthier, Jalen A. Smith, Jamal El Saghir, Sean Eddy, Mathew Alaba, Samantha Wheeler, Virginia Vega-Warner, Bradley Godfrey, Fadhl Alakwaa, Maria Larkina, Felix Eichinger, Rajasree Menon, Akihiro Minakawa, Matthias Kretzler, Shinuo Weng, Ann L. Miller, Jennifer L. Harder

## Abstract

Poor outcomes in proteinuric kidney diseases are challenging to successfully manage therapeutically due to the heterogeneity of underlying disease pathogenesis and associated risk for progression. The role of cytoskeleton-associated proteins, including the scaffolding protein Anillin (ANLN), are of specific interest in kidney disease given the importance of actin dynamics in the kidney’s specialized epithelial cell types. In this study, we identify the prevalence of genetic variants in *ANLN*, the gene encoding ANLN, in a cohort of deeply phenotyped individuals with non-diabetic proteinuric kidney disease. Thirty-one individuals (of 864 genotyped) harbor heterozygously expressed variants in *ANLN*; 7 unrelated individuals shared the same variant (I1109V) in the C-terminal pleckstrin homology (PH) domain, a region necessary for interaction with the plasma membrane. Kidney organoids generated from I1109V induced pluripotent stem cells from 1 of these individuals showed increased epithelial cell mitogen-activated protein kinase 8 network activity and apoptosis, which was enhanced by tumor necrosis factor alpha (TNF-α) and phenocopied by actin polymerization inhibition. TNF-α-treated I1109V organoids also exhibited tubular lumen expansion. Knockdown and re-expression of the analogous ANLN variant in *Xenopus laevis* embryonic epithelia resulted in defects in cell-cell junction dynamics including wavy cell membranes exhibiting increased transverse movements as well as abnormal junctional F-actin remodeling in response to mechanical stress and leaky barrier function. Taken together, these results indicate that enhanced tubular epithelial cell death, perturbed cell-cell contacts and barrier function defects are associated with a novel ANLN variant discovered in individuals with non-diabetic proteinuric kidney disease.

**ONE SENTENCE SUMMARY:** Enhanced tubular epithelial cell death and perturbed cell-cell junction integrity and barrier function are associated with a novel Anillin coding variant discovered in a cohort of individuals with proteinuric kidney disease.

## INTRODUCTION

Proteinuric kidney disease (ProtKD) encompasses diseases of various pathomechanisms that affect the glomerular filtration surface of the kidney nephron and is associated with an increased risk of progression to end stage kidney disease. Genome-wide association studies have identified genetic risk factors associated with kidney disease progression for diabetic and chronic kidney disease (*1, 2*). However, rare diseases associated with ProtKD including focal segmental glomerular sclerosis (FSGS), minimal change disease (MCD), IgA nephropathy (IgAN) and membranous nephropathy (MN) are not well represented in these studies. Apart from a few recent successes (*3, 4*), treatment strategies for these diseases remain stubbornly non-specific and of limited success. In FSGS, diverse factors contribute to significant heterogeneity of disease including variants in structural genes, genetic risk alleles, infection, pregnancy and drug exposures, suggesting that specific insults may cause disease in genetically pre-disposed individuals (*5, 6*). This disease heterogeneity exemplifies both the challenge for successful treatment design and the potential benefit of personalized therapy based on an individual’s disease characteristics.

NEPTUNE (Nephrotic Syndrome Study Network) was designed to molecularly phenotype rare ProtKD based on a combination of longitudinal clinical, genetic, biosample and kidney tissue morphologic data (*7*). NEPTUNE includes data from over 800 adult and pediatric participants without previously identified monogenic causes for their diseases drawn from over 30 study sites in North America. Investigations of this cohort have yielded critical insights into the heterogeneous pathophysiology of FSGS and MCD, including identification of tumor necrosis factor alpha (TNF-α) pathway activation in kidney tissue in a subset of individuals whose kidney disease progressed (*8*). As maintenance of epithelial barrier function in the intestine following TNF-α enhanced apoptosis and cell shedding requires a dynamic cytoskeleton (*9*), we posited that coding variants of cytoskeleton-associated genes could impair kidney epithelial cell response to stressors. Based on prior descriptions of 3 familial FSGS kindreds associated with mutations in ANLN (*10–12*), here we queried and identified individuals in NEPTUNE harboring coding variants in *ANLN*.

ANLN is a scaffolding protein that tethers cytoskeletal elements to the plasma membrane, has an essential role in cytokinesis, is expressed widely in epithelial cell types and is well-conserved across vertebrates (*13–18*). ANLN localizes to podocyte foot processes at the slit diaphragm which are tight junction-like structures, where it is critical for glomerular filtration and barrier function (*10*). In contrast, ANLN’s function in kidney tubular epithelial cells has not been evaluated. Previous live cell imaging studies in *Xenopus laevis* (*X. laevis*) embryonic epithelia revealed that knockdown or overexpression of ANLN caused alterations in epithelial cell membrane morphology, defects in cell-cell junctions and barrier function and abnormal accumulation of actomyosin networks (*19–21*). In this study, we used both induced pluripotent stem cell (iPSC)-derived human kidney organoids and *X. laevis* embryonic epithelia to test the hypothesis that a novel coding variant of ANLN associated with ProtKD could impact tubular epithelial cell structural stability resulting in altered cell resilience under stress.

## RESULTS

### *ANLN* coding variants are detected in individuals with proteinuric kidney disease

A targeted search for genetic variants potentially impacting the function of ANLN was performed in NEPTUNE study participants. A total of 26 individuals with 15 unique heterozygously-encoded *ANLN* variants within the coding regions were identified (Fig. 1A-B). Two additional variants outside the coding regions of *ANLN* were identified in 5 individuals (Fig. S1A). None of these variants has been previously described in association with ProtKD. Three previously characterized ANLN coding variants associated with FSGS (*10–12*) were not identified among the NEPTUNE study participants. Coding variants affecting the C-terminal domains of ANLN were more common; 65.4% of all variants were in the RNA binding domain (RBD), C2 and PH domains combined, despite these regions comprising 36.7% of the total protein coding region (Fig. 1A). Further, 9 individuals (34.6%) had variants that localized to the PH domain (comprising 14% of total protein length) and 7 of those individuals all had the same single missense mutation of an isoleucine to valine at position 1109 (allele frequency 0.0081) (Fig. 1B). The frequency of this I1109V variant was double the observed population frequency (gnomAD allele frequency 0.00358). Further, we searched for additional genetic variants in the 7 individuals with the I1109V variant based on known monogenic contributors to ProtKD and cytoskeleton-related pathways (Table S1-S2); a small number of additional heterozygous coding variants were identified, but none were detected in more than one individual or known to be associated with pathogenicity (Fig. S1B). Thus, our analysis identified multiple coding variants enriched in the C-terminal region in ANLN in a cohort of individuals with ProtKD. One particular variant within the PH domain, I1109V, was overrepresented in the cohort, suggesting a role for this variant in the pathogenesis of ProtKD.

**Figure 1.**
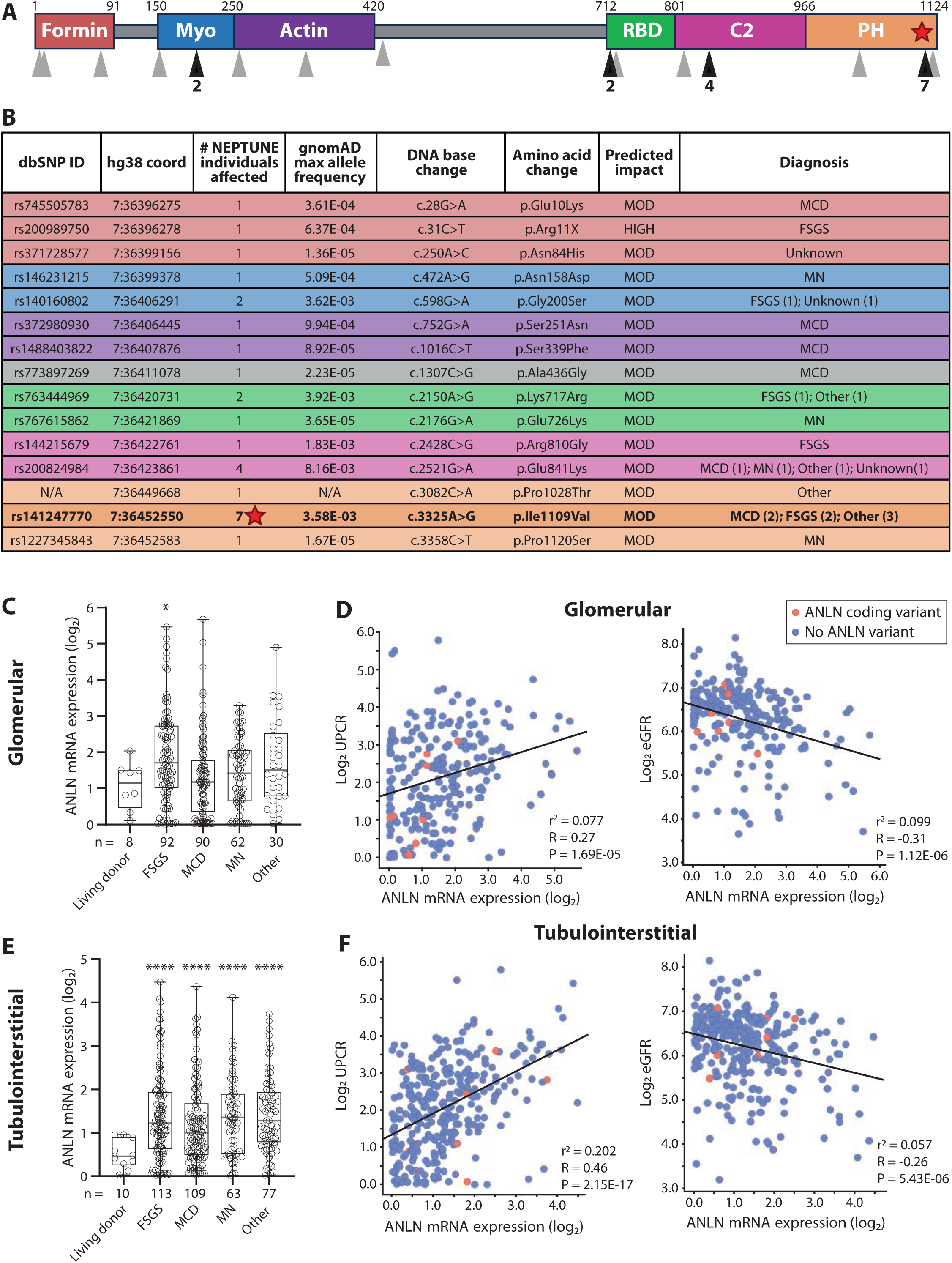
A*NLN* coding variants identified in NEPTUNE study participants. **(A)** ANLN protein domain diagram. Arrowheads indicate coding variants listed in (B) (Black: variant in multiple individuals. Gray: variant in one individual). **(B)** ANLN coding variants identified in NEPTUNE study participants. Rows are colored according to protein domains in (A). “Other”: IgAN, Diabetic nephropathy, Membranoproliferative glomerulonephritis (MPGN), Immune complex glomerulonephritis. “Unknown”: not biopsied, withdrew/not biopsied, ineligible biopsy. **(C and E)** Box plots showing glomerular (C) and tubulointerstitial (E) expression of ANLN in individuals with proteinuric kidney diseases FSGS, MCD and MN compared to healthy living transplant donors. **(D and F)** Scatter plots showing correlation of ANLN mRNA expression with UPCR (left plots: n = 238 (D); n = 306 (F)) and eGFR (right plots: n = 232 (D); n = 298 (F)) in glomerular (D) versus tubulointerstitial (F) tissue from NEPTUNE participants. (A) and (B): Red star highlights I1109V variant present in 7 unrelated individuals. (C) and (E): box plots depict median, 25^th^ and 75^th^ percentiles and min/max. \**p*<0.05, \*\*\*\**p*<0.0001, two-tailed t-test. (D) and (F): magenta points indicate individuals with an ANLN coding variant. Simple linear regression used.

### *ANLN* expression is elevated in proteinuric kidney disease

To further explore alterations of *ANLN* expression in ProtKD, we compared transcript levels in kidney tissue from individuals with ProtKD from the NEPTUNE cohort to those with healthy kidneys. *ANLN* mRNA abundance was significantly increased in glomerular tissue from individuals with FSGS (Fig. 1C) and correlated positively with urine protein loss (urine protein creatinine ratio, UPCR) and negatively with kidney function (estimated glomerular filtration rate, eGFR) (Fig. 1D). *ANLN* mRNA abundance was also significantly increased in tubulointerstitial tissue from individuals with FSGS, MCD and MN (Fig. 1E) and associated positively with UPCR and negatively with eGFR (Fig. 1F) as was seen in glomerular tissue. Moreover, *ANLN* mRNA levels correlated with a composite expression score of TNF pathway activation (Fig. S1C-D), which we previously showed as being associated with poor outcome in FSGS and MCD (*8*). Further, *ANLN* mRNA expression was similarly increased in kidney tissue from individuals in another kidney disease cohort from the European Renal cDNA Bank (ERCB) (*22*), in glomerular tissue from individuals with MCD (Fig. S1E) and in tubulointerstitial tissue from individuals with FSGS and MCD (Fig. S1F). Previous immunohistochemical staining of kidney tissue from an individual with FSGS revealed enhanced ANLN protein expression in glomeruli (*10*) providing evidence that ANLN levels are also affected in disease. Together, these results revealed perturbed *ANLN* mRNA expression in both glomerular and tubulointerstitial compartments in ProtKD, suggesting that altered *ANLN* mRNA expression is associated with kidney epithelial cell dysfunction.

### *MAPK8* transcriptional network activity is enhanced in I1109V-iPSC kidney organoid epithelial cells

We next explored a potential pathogenic effect of I1109V on epithelial cell function. The C-terminal PH domain Ile1109 residue (Fig. 1A-B; red star) is highly conserved across species (Fig. 2A), suggesting a key role for this residue. While the N-terminal domains of ANLN interact with actomyosin, the C-terminal domains facilitate interaction with the plasma membrane. The latter is mediated by association with phosphatidylinositol 4,5-bisphosphate (PIP_2_) in lipid rafts which supports ANLN-mediated localization of active RhoA at the plasma membrane (Fig. 1A and Fig. 2B) (*14, 15, 17, 18*). Thus, we hypothesized that I1109V would destabilize the ANLN-mediated interaction of the cytoskeleton with the plasma membrane. To test this hypothesis, we utilized kidney organoids generated from an iPSC line derived from 1 of the 7 individuals harboring the I1109V variant (Fig. 2C). Our experimental strategy is shown in Fig. 2D. I1109V kidney organoids developed well-structured epithelialized tubules as well as podocytes and stromal cells (Fig. 2E and Fig. S2A). Moreover, I1109V kidney organoids expressed similar levels of collagen (COL1) and fibronectin (FN1) to wild-type (WT) organoids, indicating that the variant organoids were not more fibrosed than WT organoids (Fig. S2B-C). However, single cell transcriptional profiling (Fig. 2F, Fig. S2D and Data File S1) of I1109V organoid tubular epithelial cells and podocytes revealed enhanced *MAPK8* (mitogen-activated protein kinase 8) network activity compared to WT cells at baseline (Fig. 2G, Fig. S2E and Data File S1).

**Figure 2.**
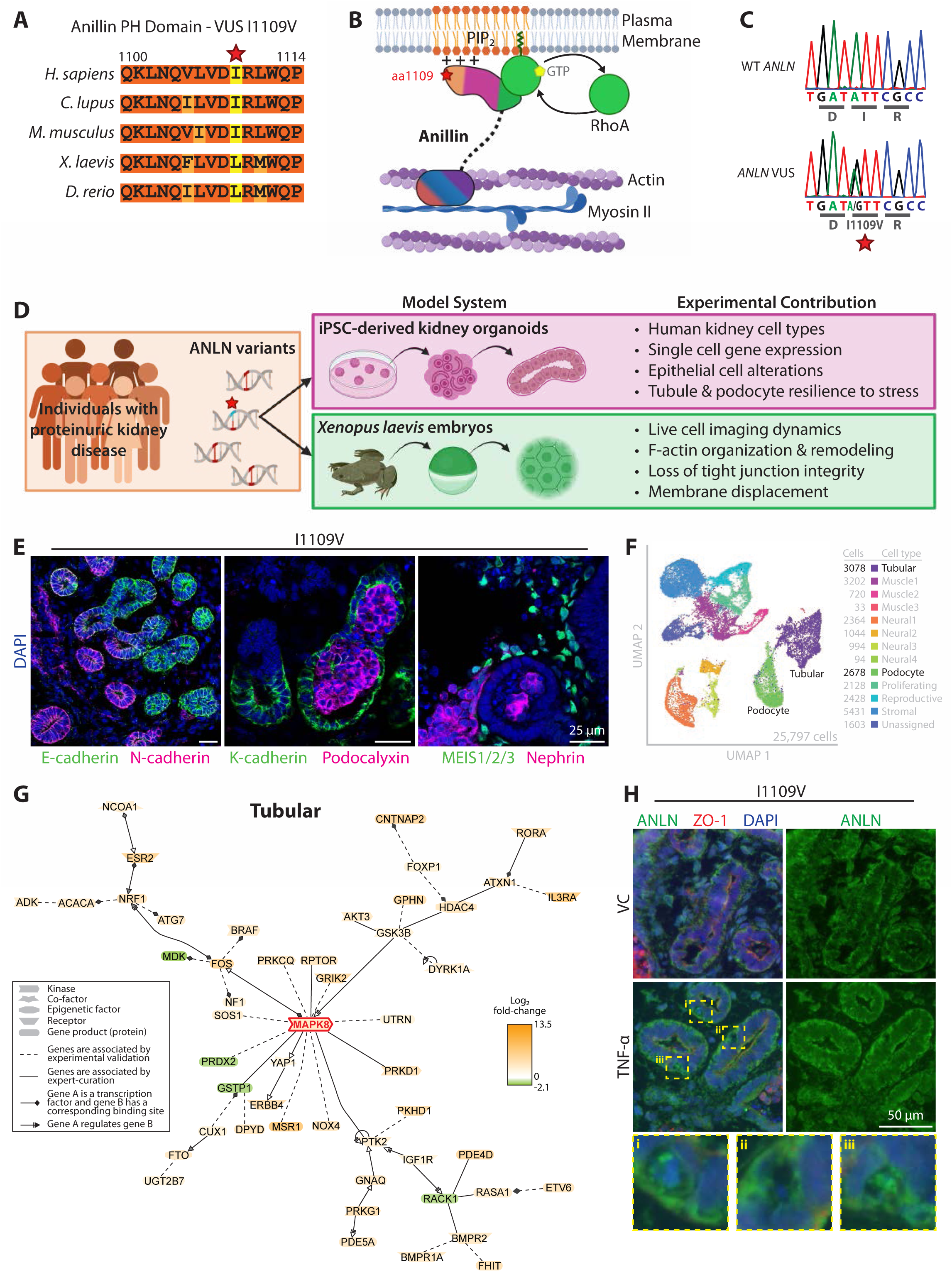
ANLN I1109V kidney organoids develop normally despite enhanced *MAPK8* pathway expression in tubular cells. **(A)** Amino acid conservation surrounding position 1109 (yellow) in the PH domain of ANLN across species. **(B)** Diagram of ANLN scaffolding cellular components. ANLN has three N-terminal domains (formin-binding [red]; myosin-binding [blue]; actin-binding [purple]) and transiently interacts with active RhoA [green]. ANLN associates with the plasma membrane via its C2 [magenta] and PH [orange] domains. Modified from Sun *et al.* (*15*). **(C)** Sanger sequencing shows missense heterozygous mutation in exon 24, 3325A>G (I1109V) in *ANLN* from NEPTUNE participant-derived iPSCs (bottom) compared to wild-type cells (top). **(D)** Methodological overview of this study. **(E)** Immunofluorescence (IF) imaging of kidney organoids generated from I1109V iPSCs showing markers of tubular cells (E-/N-/K-cadherin), podocytes (podocalyxin, nephrin) and stromal cells (MEIS1/2/3). **(F)** Uniform Manifold Approximation and Projection (UMAP) representation of single cell transcriptomes of D25 I1109V and D24 wild-type untreated organoids (12 cell type clusters). **(G)** Literature-based network analysis from the top 500 differentially regulated genes (FDR<0.01, Abs Log2 fold-change≥1.0) in I1109V variant epithelial (tubular) cells compared to wild-type. GePS literature-based network represents the top 50 best connections. **(H)** IF imaging of ANLN expression in I1109V kidney organoids after TNF-α treatment (VC: PBS vehicle control). Dashed boxes highlight areas with abnormal staining.

TNF-α can activate MAPK8 and induce actin cytoskeletal rearrangements in glomerular epithelial cells (*23, 24*). Moreover, TNF-α pathway activation is associated with worse outcomes in individuals diagnosed with MCD or FSGS (*8*). Thus, we employed our previously established TNF-α stressed organoid model of kidney disease to explore TNF-α effects on I1109V organoid epithelial cells (*25*). TNF-α-treated I1109V kidney organoids exhibited the expected increase in expression of the pro-inflammatory cytokine *CXCL10* (Fig. S3A). ANLN immunostaining increased throughout the cross-section of tubular cells in I1109V kidney organoids following TNF-α treatment (Fig. 2H); while visually striking, quantification did not reach significance (*p=*0.0906) (Fig. S3B). We noted areas within the TNF-α-treated tubules that displayed increased frequency of abnormal ANLN staining pattern compared to control (Fig. 2H, yellow boxes). Super-resolution microscopy imaging of TNF-α-treated tubules revealed instances of tubular cells with concentrated F-actin staining at the cell periphery along with concentrated, punctate nuclear staining, suggesting apoptotic cells (Fig. S3C bottom, cyan arrowheads). Our imaging results also suggested that TNF-α treatment increased waviness and prominence of cortical F-actin at I1109V tubular cell-cell junctions (stained with Zonula Occludens-1 [ZO-1]) (Fig. S3C). However, imaging analysis was hampered by the 3-dimensional organoid tubular structure. These results demonstrate that while I1109V organoid epithelial cells appear structurally normal, the MAPK8 pathway is already activated at baseline, and TNF-α treatment appeared to alter tubular epithelial cell structure.

### ANLN I1109V tubular epithelial cells are more susceptible to injury

To further investigate TNF-α-mediated I1109V organoid epithelial cell structure changes, we stained for apoptotic cells. In TNF-α-treated I1109V kidney organoids, numerous cleaved caspase 3 (cC3) positive apoptotic tubular cells were observed throughout tubule structures with some appearing to be extruding into the lumen (Fig. 3A, yellow arrows). Additionally, apoptotic cells and cell debris accumulated in the lumen, coupled with a loss of columnar tubular cell height (Fig. 3B), reminiscent of a partial epithelial-to-mesenchymal transition (EMT) (*26, 27*). Since partial EMT would be expected to result in a proportionally larger lumen area relative to the entire tubule area as a result of loss of columnar cell height (Fig. 3C), the percentage of total tubule area occupied by tubular cells compared to lumen area was quantified. Tubules in I1109V kidney organoids had proportionally larger lumens following TNF-α treatment, whereas WT organoids treated with TNF-α did not display altered tubule proportions (Fig. 3D). To ensure that a difference in lumen size did not reflect a difference in proximal and distal tubular lumen morphology, lumen proportions for both regions were measured and compared. Indeed, I1109V lumen size increased following TNF-α treatment in both proximal and distal tubular regions (Fig. S3D-E). Further, TNF-α enhanced tubular cell transcriptional expression of key EMT cellular markers in I1109V kidney organoids (Fig. 3E, Fig. S4A-B and Data File S1).

**Figure 3.**
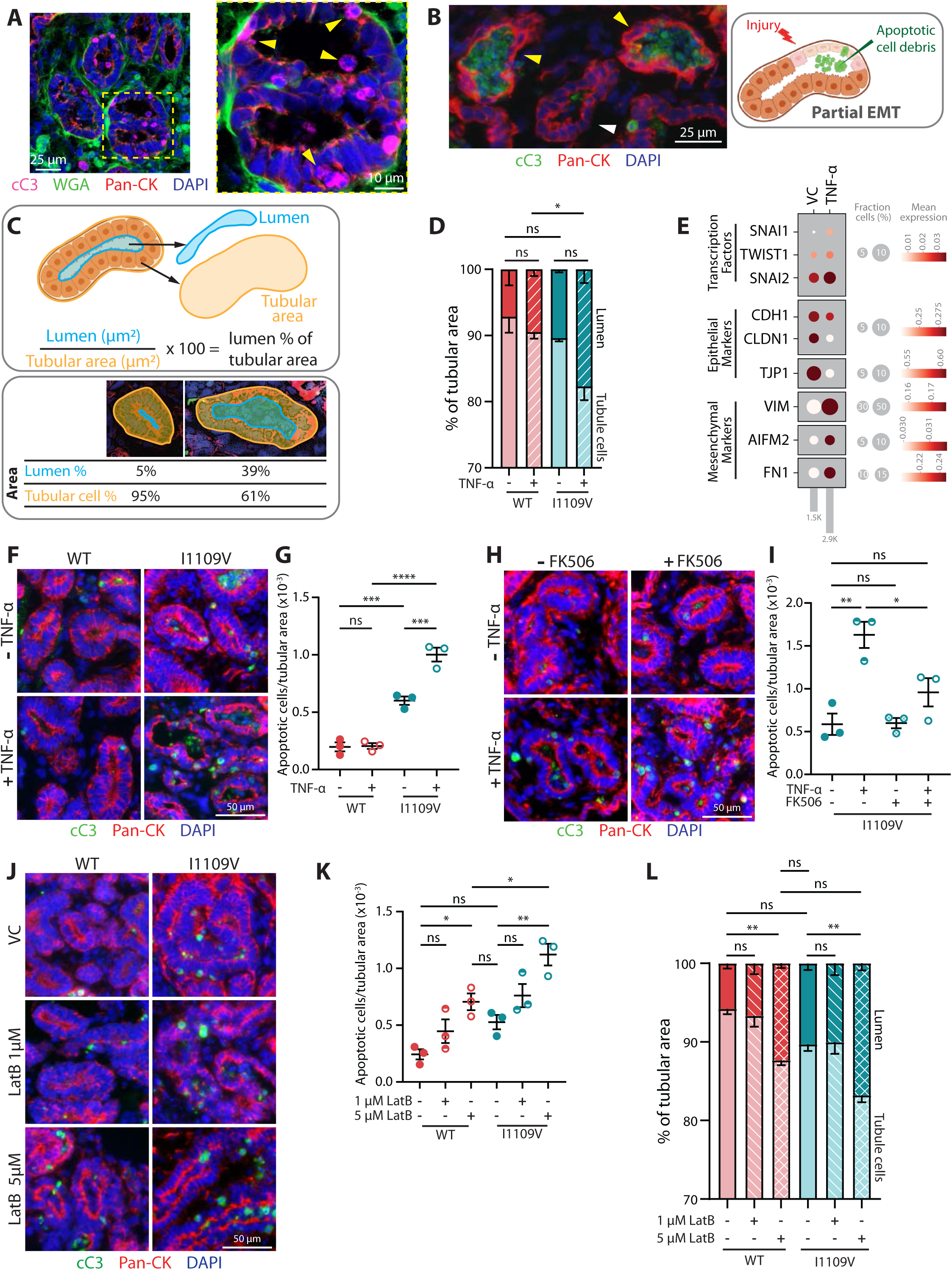
I1109V kidney organoids are more susceptible to tubular cell disruption and apoptosis. **(A)** Sum Z-projections of confocal IF imaging of cleaved caspase-3 (cC3) in I1109V organoid tubular cells (pan-cytokeratin [Pan-CK]) after TNF-α treatment. Dashed box indicates higher magnification area highlighting apoptotic cells (right, yellow arrowheads). **(B)** Left: IF imaging of cC3 expression in TNF-α-treated I1109V organoid displays injured tubules (yellow arrowheads) compared to a healthy tubule (white arrowhead). Right: diagram illustrates kidney tubular cell injury resulting in partial EMT and apoptotic cellular debris in the lumen. **(C)** Top: schematic of kidney organoid tubule quantification: tubule and lumen perimeters were outlined (orange and blue, respectively) based on Pan-CK. Bottom: representative tubules showing a higher percentage of lumen to tubular area (right) suggesting tubular cell collapse and EMT. **(D and L)** Stacked bar graph shows percentage of lumen (darker color) and tubule (lighter color) area in WT and I1109V organoids after TNF-α (D) or LatB (L) treatment. **(E)** Dot plot showing EMT-related gene expression in tubular cell clusters, comparing untreated and TNF-α-treated I1109V kidney organoids. **(F, H and J)** IF imaging of cC3 in WT and I1109V kidney organoid tubular cells (Pan-CK) treated with TNF-α (F); TNF-α and/or FK506 (H); and LatB (J). **(G, I and K)** Quantification of apoptotic cells per tubule area based on IF staining (F, H, J, respectively). Data for (D), (G), (I), (K) and (L) are displayed as means +/- SEM (n = 3) using one-way ANOVA with Šídák’s test. All comparisons made are shown. \**p*≤0.05, \*\**p*≤0.01, \*\*\**p*≤0.001, \*\*\*\**p*≤0.0001.

The difference in apoptotic cell fraction of TNF-α-treated I1109V versus WT kidney organoid tubular cells and podocytes was then assessed. I1109V tubular cells displayed a significantly higher baseline of apoptosis compared to WT. TNF-α treatment further enhanced I1109V tubular cell apoptosis, while WT cells were unaffected (Fig. 3F-G). TNF-α treatment elicited a slight increase in podocyte apoptosis, though no discernable difference between WT and I1109V podocytes was appreciated (Fig. S4C-E). Together, these data suggest that I1109V tubular cells are more susceptible to TNF-α-mediated cellular injury, and we propose this prompts remaining tubular cells to compensate and attempt to rescue the tubular structure.

We next aimed to alleviate TNF-α-mediated tubular cell injury in I1109V organoids by interfering with the background activation of the MAPK8 pathway. To do this, we used FK506 (tacrolimus), a calcineurin inhibitor therapeutically used in ProtKD (*28*), which can suppress MAPK8 pathway activation independent of its calcineurin inhibitory effects (*29, 30*). Additionally, FK506 increases expression of FKBP12/FKBP1A which binds to and stabilizes F-actin in podocytes (*31*) and suppresses cofilin-mediated F-actin destabilization in neuronal dendrites (*32*). Thus, we hypothesized that FK506 would mitigate epithelial cell injury susceptibility associated with ANLN I1109V. Indeed, apoptosis was decreased in I1109V tubules treated with FK506 in combination with TNF-α compared to TNF-α treatment alone (Fig. 3H-I). However, FK506 alone did not alter the higher level of baseline apoptosis in I1109V tubular cells. Tubular cells in WT kidney organoids were unaffected by FK506 treatment in the presence and absence of TNF-α (Fig. S4F-G). These results suggest that calcineurin inhibition may help modulate TNF-α-mediated stress in ANLN I1109V variant tubular cells, though no substantial benefit was apparent for unstressed I1109V cells despite baseline enhanced *MAPK8* transcriptional network activity.

Since ANLN functions as a scaffold for multiple cytoskeletal components, we hypothesized that I1109V tubular cells would show enhanced sensitivity to actin destabilization. Indeed, inhibiting actin polymerization in I1109V organoids with latrunculin B (LatB) caused an increase in tubular cell apoptosis (Fig. 3J-K). High LatB dosing (5 µM) resulted in near-total loss of F-actin, though ANLN expression and localization appeared unaffected (Fig. S5A). In contrast, WT kidney organoids showed a modest increase in baseline tubular cell apoptosis, with apoptosis at high LatB concentration similar to levels seen in untreated or minimally treated I1109V cells (Fig. 3K). In addition to F-actin destabilization phenocopying tubular cell apoptosis from TNF-α treatment, LatB similarly caused proportionally larger tubular lumens in I1109V kidney organoids (Fig. 3L). Conversely, treatment with the actin stabilizer jasplakinolide (JAS) had no impact on tubular cell apoptosis in I1109V organoids (Fig. S5B-C). These results suggest that I1109V’s increased tubular cell sensitivity to destabilization of actin may be mediated by impairment of ANLN’s scaffolding function that is especially apparent during periods of active actin cytoskeleton remodeling, in contrast to periods of relative cytoskeletal stability.

Thus far, our evaluation of I1109V’s function in human kidney organoid epithelial cells has relied on event capture and analysis using a static organoid system. This approach has distinct advantages from the perspective of disease modeling including using iPSCs from an individual with the disease in a model with architecture incorporating multiple kidney cell types that are difficult to generate in other systems. Indeed, our analysis revealed that the I1109V ANLN variant altered *MAPK8* transcriptional network activation in multiple epithelial cell types and enhanced tubular cell susceptibility to stressors accompanied by tubular cell morphology changes suggestive of EMT. However, to further investigate I1109V’s mechanisms contributing to enhanced cell injury, we opted to employ a well-established model system for live cell imaging of epithelial cell-cell junction and cytoskeletal dynamics: *X. laevis* (African clawed frog) embryos (Fig. 2D).

### *X. laevis* embryos expressing ANLN 1109V are susceptible to junction separation under mechanical stress

The embryonic epithelium of *X. laevis* embryos is an ideal model to investigate ANLN, as there is high degree of conservation between the human and *X. laevis* Anillin protein sequences, particularly in the PH domain (Fig. S6A-C) (please note that we will also refer to the *X. laevis* Anillin protein as ANLN for simplicity). The isoleucine (I) at position 1109 is well conserved across species (Fig. 2A); in *X. laevis*, it is a leucine (L). Therefore, we generated the analogous variant, which we refer to as ANLN 1109V.

We knocked down (KD) endogenous ANLN in the embryos using a validated anti-sense morpholino (Fig. S6D), reducing the ANLN protein level by about 50% (Fig. S6E) (*19–21*). Bulk RNAseq was performed on WT and ANLN KD embryos to examine whether altered ANLN protein levels affect similar cellular transcriptional pathways in *X. laevis* to those modified in human kidney cells. Principal component analysis revealed reasonable clustering of WT and KD samples (Fig. S7A). Expression of *anln* mRNA was more variable in the KD sample compared to WT (Fig. S7B). As morpholino KD of ANLN blocks translation, not transcription, it is likely that the variability in *anln* mRNA expression reflects a cellular response to decreasing ANLN protein levels (Fig. S6E). We next mapped the *X. laevis* reads to human orthologs to evaluate pathways affected by ANLN KD. Several pathways involved with cytoskeletal component binding and activity were among the most highly activated gene pathways (Fig. S7C and Data File S1), suggesting that ANLN KD may result in a compensatory increase in expression of other cytoskeletal factors to maintain cellular homeostasis. By demonstrating cytoskeletal pathway transcriptional effects of ANLN alterations shared by human and *X. laevis*, this analysis supports our use of this model to investigate the I1109V variant.

To model the heterogeneous ANLN I1109V variant discovered in our ProtKD cohort in *X. laevis* embryos, we microinjected mRNAs encoding either WT or ANLN 1109V into ANLN KD embryos. We then allowed them to develop to gastrula stage, at which point the embryos had formed a polarized epithelium with apical cell-cell junctions facing the external environment; this allows cell-cell junctions and the actin cytoskeleton to be easily viewed with confocal microscopy, while leaving the embryo completely intact. Immunostaining for ZO-1 showed comparable overall cell morphology across control, ANLN KD, ANLN KD+WT and ANLN KD+1109V embryos (Fig. S8A). ANLN KD embryos exhibited decreased ZO-1 intensity, as expected (*19–21*), while ANLN KD+1109V embryos often showed decreased or non-uniform ZO-1 intensity at junctions (Fig. S8A), though the effect was variable. Notably, unlike the sharp, linear junctions of ANLN KD+WT embryos, junctions in ANLN KD+1109V embryos often appeared ‘separated’ following fixation, immunostaining and mounting for microscopy, during which mechanical stress is applied to the tissue (Fig. 4A). Indeed, we quantified a significant increase in junction separation (two distinct lines of ZO-1 intensity) in ANLN KD+1109V embryos compared with either ANLN KD+WT or control embryos (Fig. 4B and Fig. S8B). Further, analysis of regions with mosaic expression – where cells that received ANLN KD+1109V are mixed with control cells – showed that junctions where at least one of the cells contributing to a cell-cell junction expressed ANLN 1109V often separated, whereas internal control cells had intact linear junctions (Fig. 4C). Together, these results suggest that expression of the ANLN 1109V variant in an ANLN KD background leads to weakened cell-cell junctions that are more susceptible to mechanical damage resulting in junction separation.

**Figure 4.**
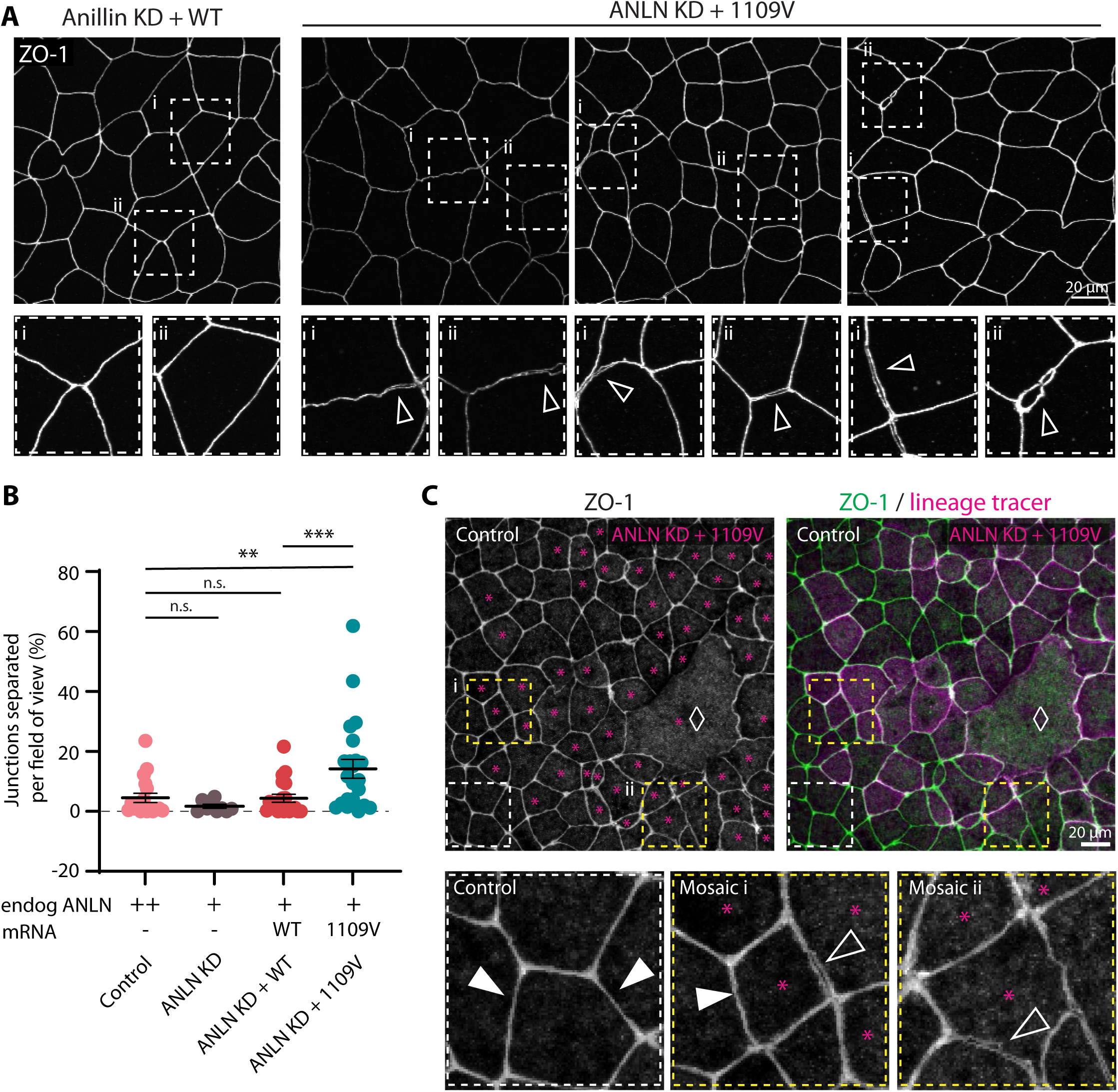
Expression of the ANLN 1109V variant in *X. laevis* embryos leads to junction separation upon fixation and mounting. **(A)** IF imaging of ZO-1 in ANLN KD+WT and ANLN KD+1109V *X. laevis* embryos. Dashed boxes highlight areas of junction separation (open white arrowheads) in ANLN KD+1109V embryos. **(B)** Quantification of junction separation determined as the percentage of separated junctions with respect to the total number of junctions in each field of view. Each data point represents a unique embryo (n = 3 experiments, 18 control (water-injected) embryos, 9 ANLN KD embryos, 20 ANLN KD+WT embryos and 23 ANLN KD+1109V embryos). **(C)** IF imaging of a mosaically-injected *X. laevis* embryo where some of the cells express ANLN KD+1109V (lineage tracer, magenta asterisks) and some of the cells are controls. Dashed white box indicates a control region, while dashed yellow boxes indicate mosaic regions highlighting junction separations (open white arrowheads). Diamond symbol indicates an abnormally large ANLN KD+1109V cell. Data for (B) are displayed as mean +/- SEM. Statistical analysis: one-way ANOVA to determine if there was a difference between control, ANLN KD, ANLN KD+WT and ANLN KD+1109V (*p* = 0.0015); Student’s t-test for pairwise comparisons between WT and ANLN KD +1109V (p = 0.0159), and between ANLN KD + WT and ANLN KD + 1190V (p = 0.0097); \*\**p*≤0.01, \*\*\**p*≤0.001.

### Expression of ANLN 1109V alters junction morphology and dynamics in *X. laevis* embryos

Live confocal microscopy of ANLN KD+WT and ANLN KD+1109V embryos expressing tagged ZO-1 (BFP-ZO-1) and a probe for F-actin (LifeAct-RFP) revealed that ANLN KD+1109V cell-cell junctions often appeared to bulge or look wavy in comparison with linear junctions in ANLN KD+WT embryos (Fig. 5A). Comparing the ‘junction linearity index’ demonstrated that ANLN KD+1109V junctions were significantly less straight than ANLN KD+WT junctions (Fig. 5B-C). Examining time-lapse movies to compare junction dynamics showed that ANLN KD+WT cell-cell junctions maintained their overall linearity over time (Fig. 5D). In contrast, abnormal junction dynamics were regularly observed in ANLN KD+1109V embryos (Fig. 5E-F). In some cases, these junctions appeared bulged, retaining their curvature over time (Fig. 5E), while in other cases they developed an increasingly wavy pattern (Fig. 5F).

**Figure 5.**
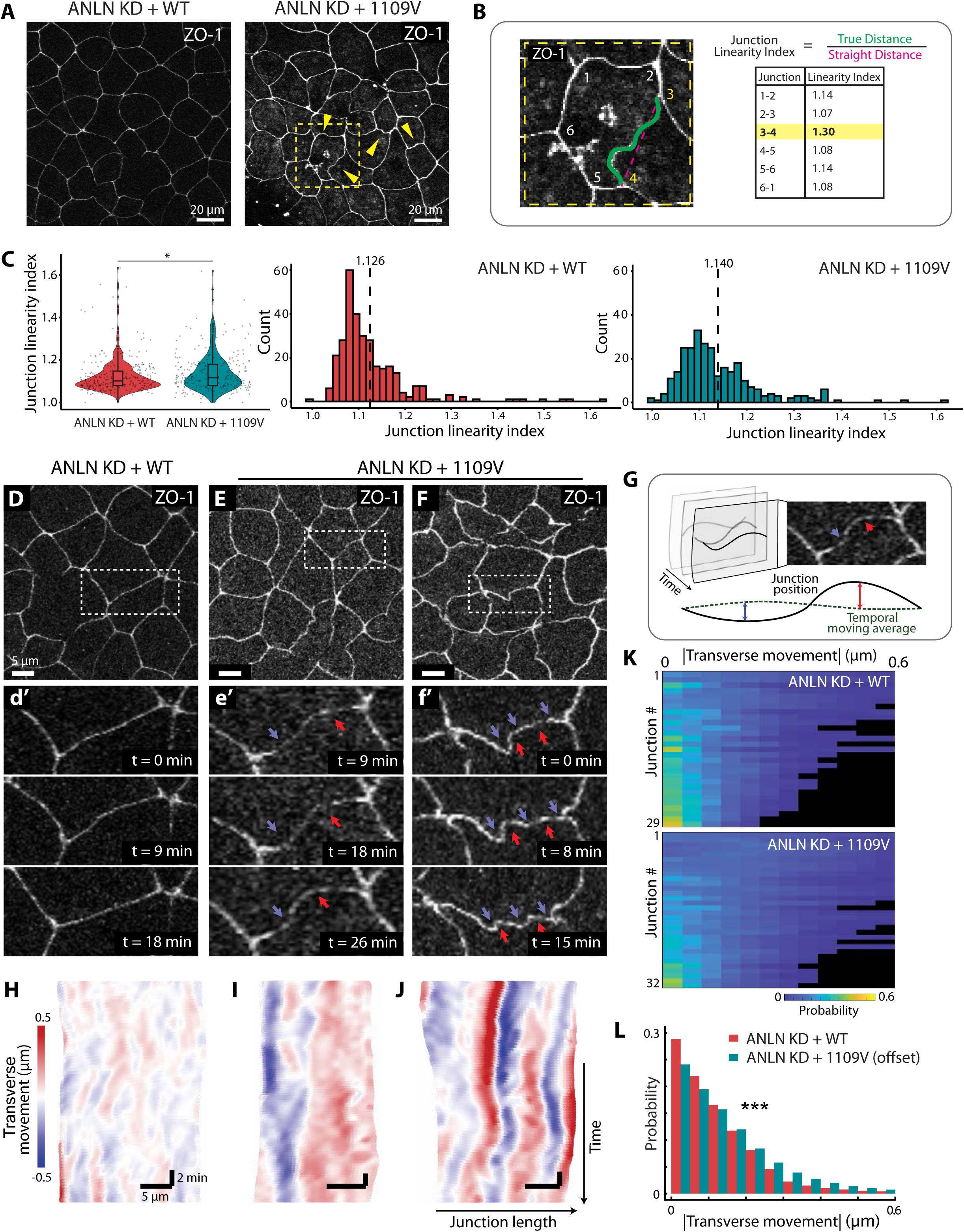
Expression of ANLN 1109V alters junction morphology and junction fluctuation dynamics in *X. laevis* embryos. **(A)** Live confocal images of ZO-1 (BFP-ZO-1) in ANLN KD+WT or ANLN KD+1109V *X. laevis* embryos. Yellow arrowheads indicate junction bulges or waviness. Dashed box highlights region enlarged in (B). **(B)** Schematic of junction linearity index and example junction linearity index values. Junctions that are straight have an index near 1, while junctions that have bulges or appear wavy have an index above 1. **(C)** Quantification of junction linearity index represented as a violin plot (left) or histograms with mean values indicated (right). n = 4 experiments, 309 ANLN KD+WT junctions, 55 cells, 9 embryos; 281 ANLN KD+1109V junctions, 54 cells, 9 embryos. **(D-F)** Representative ZO-1-labeled cells in ANLN KD+WT or ANLN KD+1109V embryos. (d’-f’) time-lapse stills showing the dynamic changes of junction linearity over time. Arrows point to increased junction bulges or waviness. **(G)** Schematic of TFlux analysis. Transverse movement is quantified as the distance between the real-time junction position and its temporal moving average. **(H-J)** TFlux kymographs of boxed junctions in (D–F). **(K)** Heat map of absolute transverse movement across junctions. n = 29 ANLN KD+WT junctions, 32 ANLN KD+1109V junctions. **(L)** Bar plots showing the pooled distribution of absolute transverse movement from all junctions analyzed in (K). Data for (C, left) are displayed as violin plots with overlaid box plots depicting median, 25^th^ and 75^th^ percentiles and min/max. Statistical analysis: Student’s t-test; \**p*≤0.05. (L) Statistical analysis: Kolmogorov-Smirnov test; \*\*\**p*≤0.0001.

To understand the underlying mechanisms resulting in altered junctional linearity and dynamics, we applied a previously described Transverse Fluctuation (TFlux) analysis (*33*), which quantifies the movement of junctions perpendicular to their local average position over time (Fig. 5G). In prior studies, sub-minute fluctuations exhibited spring-like alternating transverse movement, visualized as alternating blue and red regions in TFlux kymographs (*33*). ANLN KD+WT junctions showed a similar pattern, even in a longer time window (Fig. 5H), suggesting a cortical tension-dominated behavior. Surprisingly, the TFlux kymographs for ANLN KD+1109V junctions displayed distinct patterns. In bulged junctions, long-lasting, parallel blue and red bands appeared (Fig. 5I), indicating persistent transverse deformation in opposite directions (Fig. 5e ‘, arrows). Wavy junctions showed similar alternating stripes, but these were narrower and spatially confined to specific segments of the junction (Fig. 5J), corresponding to localized zigzags along the junction (Fig. 5f ‘, arrows).

To quantify the overall difference, we measured absolute transverse movement at each time point across multiple junctions. Both individual and pooled data revealed a clear and significant increase in transverse junction movement in ANLN KD+1109V embryos compared to ANLN KD+WT (Fig. 5K-L and Fig. S9). The wavy junctions and enhanced transverse fluctuations are likely a combined effect of both increased transverse forces perpendicular to junction and reduced longitudinal tension along junctions (*20, 34*). Together, these results suggest that the ANLN 1109V variant may disrupt the proper dynamic connection of the actin cytoskeleton to cell-cell junctions.

### ANLN 1109V expression in *X. laevis* embryos alters junctional F-actin remodeling during mechanical challenge

Junction morphology is dependent on robust scaffolding to the actomyosin cytoskeleton (*35–37*) by proteins including ANLN (*17, 19, 20*). Therefore, we hypothesized that the wavy junctions and abnormal dynamics observed in ANLN KD+1109V embryos could be due to changes in F-actin organization and remodeling. We analyzed F-actin dynamics at sites of junction bulging in ANLN KD+1109V embryos compared with control junctions. Indeed, we found that F-actin accumulation was imbalanced – higher on one side of the junction than the other – for bulged junctions in ANLN KD+1109V embryos compared to straight junctions in ANLN KD+WT embryos, where the actin accumulation was similar on both sides of the junction (Fig. 6A-B, Fig. S10A and Movie S1). Combined with the data presented earlier demonstrating that the I1109V mutation increases sensitivity to actin destabilization via LatB treatment (Fig. 3J-L), our results provide further evidence that the I1109V human variant affects ANLN’s ability to scaffold and properly regulate junctional F-actin, leading to abnormal cell-cell junctions.

**Figure 6.**
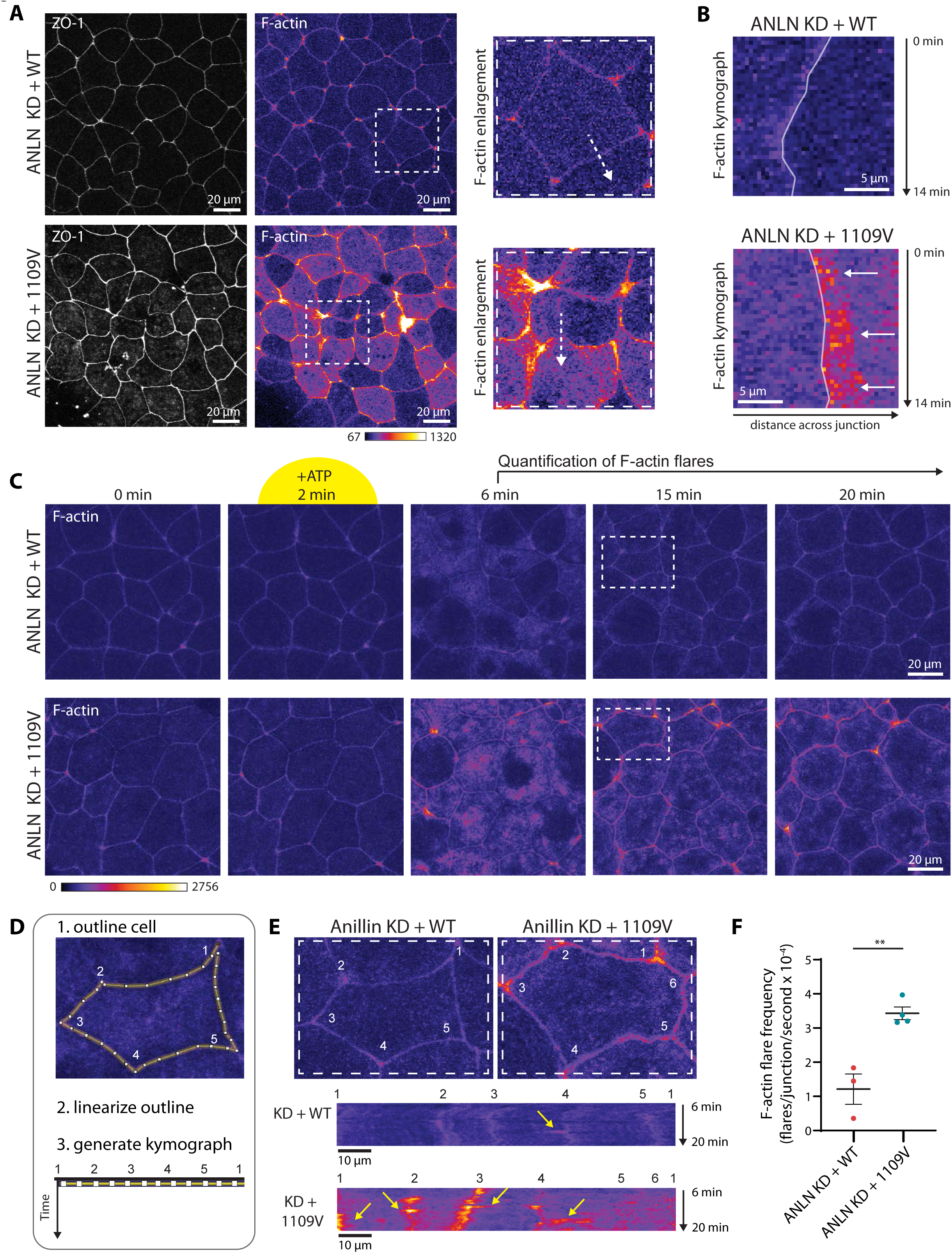
*X. laevis* embryos expressing ANLN 1109V exhibit abnormal junctional F-actin remodeling during mechanical challenge. **(A)** Live confocal images of ZO-1 (BFP-ZO-1) and F-actin (LifeAct-RFP, Fire LUT) in ANLN KD+WT or ANLN KD+1109V *X. laevis* embryos. Dashed white boxes indicate regions enlarged on the right, white arrows indicate region of kymographs in (B). **(B)** Kymographs of F-actin intensity over time (generated from dashed white arrows in (A)). Junctions are marked with lines; arrows indicate repeated F-actin accumulation on one side of the junction in ANLN KD+1109V embryo. **(C)** Time-lapse stills of F-actin in ANLN KD+WT or ANLN KD+1109V embryos before and after extracellular ATP addition (yellow highlight) to induce mechanical challenge. Quantification was carried out from 6 - 20 min. Dashed white boxes indicate regions enlarged for schematic representation in (D). **(D)** Schematic of linearized cell perimeter kymograph. **(E)** Live confocal images of F-actin (top) enlarged from the dashed white boxes in (C) used to generate kymographs (bottom); cell vertices are numbered. Stable F-actin intensity appears as a vertical stripe, whereas local, transient increases of F-actin intensity – ‘F-actin flares’ – occur on or near vertices or on bicellular junctions (yellow arrows). **(F)** F-actin flare frequence quantified from minutes 6-20 to avoid large increases in cortical F-actin contraction that happen during immediately following extracellular ATP addition (at 2 mi). Each data point represents the frequency of F-actin flares counted in one field of view. n = 3 experiments, 3 ANLN KD+WT embryos, 4 ANLN KD+1109V embryos. Data for (F) are displayed as mean +/- SEM. Statistical analysis: Student’s t-test; \*\**p*≤0.01.

*In vivo*, human kidney epithelial cells are constantly subjected to different stressors that challenge cytoskeletal and junctional integrity, including alternating hemodynamic and fluid pressures as well as cytokines. Therefore, we next examined how live embryos responded to an acute mechanical challenge: exogenous ATP addition, which causes rapid, medial-apical actomyosin contraction of cells in *X. laevis* embryos, leading to contractile forces acting on cell-cell junctions (*20, 38–41*) (Fig. S10B). When ATP is added, both ANLN KD+WT and ANLN KD+1109V exhibit the expected increase in cortical contraction (*40*). Notably, in comparison with ANLN KD+WT, ANLN KD+1109V embryos showed a marked increase in dynamic ‘F-actin flares’, which are indicative of junction remodeling processes (Fig. 6C and Movie S2) (*42*). For ease of visualization of F-actin flares, kymographs were generated that depict the perimeter of the cell as a single line over time (Fig. 6D) (*19*). In ANLN KD+WT embryos, F-actin flares were generally observed at or near tricellular junctions (Fig. 6E) (*19*). In contrast, ANLN KD+1109V embryos had an increased number of F-actin flares that frequently spread along bicellular junctions as well (Fig. 6E). Quantification confirmed a significant increase in the number of F-actin flares in ANLN KD+1109V embryos (Fig. 6F). These results indicate that the ANLN 1109V disrupts normal actin-mediated remodeling of cell-cell junctions in response to mechanical challenge. We propose that the 1109V variant may disrupt ANLN’s ability to efficiently establish the scaffolding of the actin cytoskeleton to the membrane during active cytoskeleton remodeling events.

### Expression of ANLN 1109V disrupts barrier function in the *X. laevis* epithelium

Junctional F-actin is required for the maintenance of cell-cell junctions and barrier function (*43–46*); thus, we hypothesized that barrier function might also be affected by the 1109V variant. To test barrier function, we used the Zinc-based Ultra-sensitive Microscopic Barrier Assay (ZnUMBA) (*42, 47*), which detects tight junction leaks in real time via FluoZin3 fluorescence. If tight junctions are intact, FluoZin3 fluorescence remains low; however, if tight junctions are impaired, FluoZin3 can diffuse through ‘leaks’ in the barrier, interact with Zn^2+^ in the media and its fluorescence increases dramatically (Fig. 7A). In ANLN KD+WT embryos, barrier leaks were rare and were rapidly repaired (Fig. 7B). In contrast, in ANLN KD+1109V embryos, there was a noticeable increase in junctional FluoZin3 intensity over time along with local barrier leaks (Fig. 7B; Movie S3). Quantification showed that ANLN KD+1109V embryos exhibited a significant increase in junctional Fluozin3 intensity over time compared with control embryos (Fig. 7C). These results show that expression of ANLN 1109V disrupts barrier function in an intact epithelium, indicating that there are both morphological and functional defects in cell-cell junctions when the ANLN 1109V variant is expressed.

**Figure 7.**
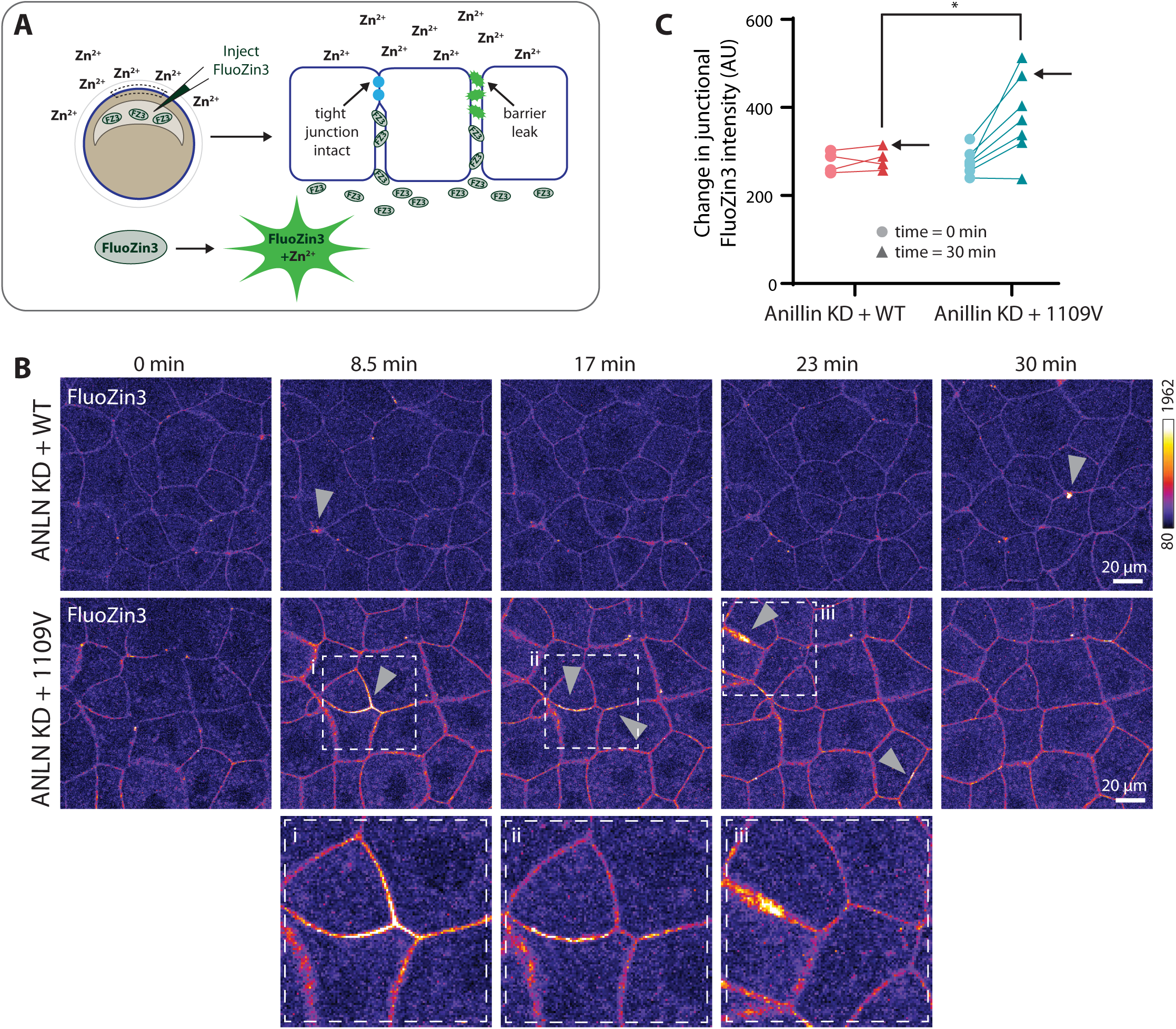
Expression of ANLN 1109V disrupts barrier function in the *X. laevis* epithelium. **(A)** Schematic of Zinc-based Ultrasensitive Microscopic Barrier Assay (ZnUMBA). **(B)** Time-lapse stills of FluoZin3 intensity (Fire LUT) in ANLN KD+WT or ANLN KD+1109V embryos. Gray arrowheads indicate barrier leaks. Dashed boxes highlight regions with barrier leaks. **(C)** Quantification of the change in junctional FluoZin3 intensity from the start of the video (time = 0 min, circles) to the end of the video (time = 30 min, triangles). Black arrows indicate values for the montages shown in (B). n = 3 experiments, 4 ANLN KD+WT embryos, 7 ANLN KD+1109V embryos. Data in (C) are paired values for same embryo at the start and end of each video. Statistical analysis: Student’s t-test, \**p*≤0.05.

In summary, our investigation of ANLN 1109V in *X. laevis* embryonic epithelial cells allowed us to model the functional impact of this human ANLN variant *in vivo* by partially knocking down endogenous ANLN and microinjecting *anln* WT or 1109V mRNA. Our data revealed that fixed embryos expressing ANLN 1109V showed susceptibility to cell-cell junction separation under the mechanical stress of mounting. The ability to do live imaging is a particular strength of this complementary system and our live imaging in *X. laevis* embryos demonstrated that ANLN 1109V expression led to perturbed cell-cell junction morphology and increased transverse fluctuation. Further, ANLN 1109V-expressing cells exhibited abnormal F-actin remodeling at junctions in response to an acute mechanical challenge, suggesting compromised actin scaffolding at cell-cell junctions. Finally, a live imaging barrier function assay revealed that expression of ANLN 1109V disrupted epithelial barrier function.

## DISCUSSION

In this study we identified a novel coding variant within ANLN’s PH domain – I1109V – in a cohort of individuals with rare forms of ProtKD. This same single residue change was found in 7 unrelated individuals at a site that is highly conserved across species, consistent with a restrictive variant. Our findings demonstrate that 1109V negatively impacts ANLN’s function as a cross-linker between the actomyosin cytoskeleton and the plasma membrane, resulting in decreased epithelial cell structural resilience to stressors, as summarized in Fig. 8.

**Figure 8.**
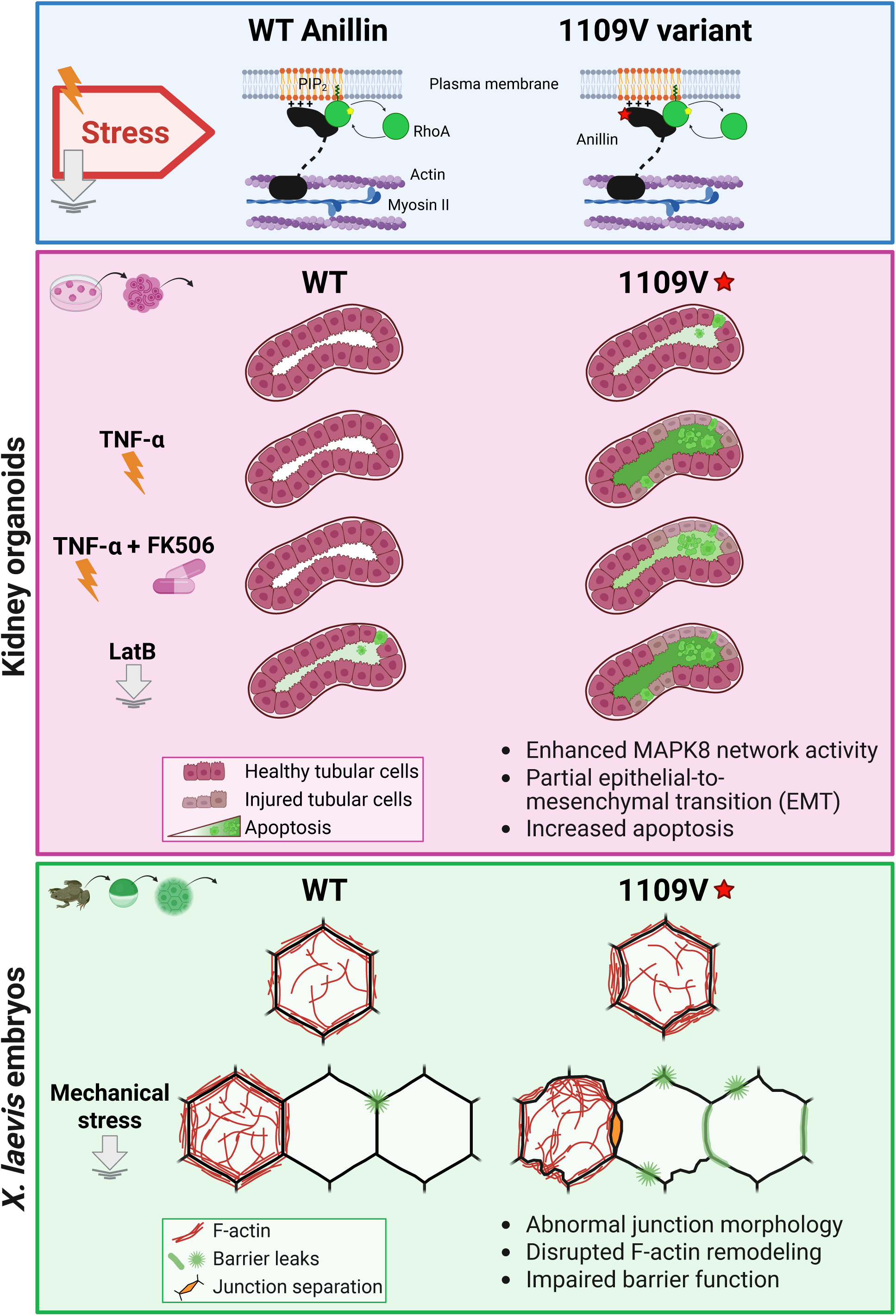
ANLN 1109V variant negatively impacts the epithelium. Summary diagram illustrating detrimental consequences of the 1109V ANLN variant in kidney organoid tubular epithelial cells and the *X. laevis* embryo epithelium.

By performing transcriptional analysis of tubular epithelial cells as well as podocytes from kidney organoids generated from iPSCs from 1 of these individuals, we showed that this variant is associated with baseline enhanced transcriptional network activity around *MAPK8*, which encodes a stress-activated kinase. Increased tubular epithelial cell injury, apoptosis and EMT provided functional evidence of cell stress and perturbed cytoskeletal dynamics at baseline, results which were compounded by exposure to cell stressors including the cytokine TNF-α and the actin destabilizer LatB. Investigation of the impact of expression of the 1109V variant in *X. laevis* embryos provided further evidence of perturbed cytoskeletal-membrane interactions including cell-cell junction fragility, abnormal actin and junctional dynamics and increased barrier leaks, some of which were further perturbed by mechanical stress. We conclude that the presence of the ANLN 1109V variant destabilizes proper actomyosin-plasma membrane interactions in epithelial cells.

However, since individuals harboring I1109V develop kidneys normally and kidney epithelial cells harboring this variant appear grossly normal, we speculate that the presence of I1109V is most impactful during periods of heightened cytoskeletal dynamics when initial actomyosin-membrane connections are occurring. Some I1109V cells fail to establish enough successful connections and succumb to apoptosis, but once cells are able to achieve a critical level of cytoskeletal scaffolding (relying on the presence of WT ANLN), perhaps additional crosslinkers bolster these connections. During periods of relative cytoskeletal stability, I1109V cells are more resilient. This interpretation aligns with our observation that FK506 does not improve baseline apoptosis frequency of I1109V in unstressed kidney tubular epithelial cells but does improve resilience to TNF-α-mediated stress.

A balance of forces at cell-cell junctions is needed to maintain tissue homeostasis. Our data suggest that when the 1109V variant is expressed, there may be an imbalance of actomyosin-based forces on junctions. When mechanical stress was applied (e.g., in this study: physical stress of mounting embryos or addition of extracellular ATP to drive acute actomyosin contraction), junctional defects including increased junction separation, changes in junction linearity and dynamics and barrier defects were observed. *In vivo*, there are many mechanical forces that can challenge junctions (e.g., cell division, morphogenesis, or physiological forces experienced by kidney cells) (*35, 48, 49*). In the future, it would be interesting to expose control and I1109V variant kidney organoids and *X. laevis* embryos to additional mechanical challenges and measure functional outputs such as barrier function.

Prior studies demonstrated the negative impact of altered ANLN levels on epithelial cell stability (*10, 11, 19–21, 50*), which aligns with our observation that increased transcriptional expression of *ANLN* is associated with proteinuria and decreased GFR. Heterogeneous expression of ANLN 1109V – even at protein levels similar to WT ANLN – affects cytoskeletal and membrane dynamics. This could indicate that there is an “effective” dose of ANLN and deviating from that can result in epithelial cell fragility, which is exacerbated by various cell stressors.

Our study demonstrates how we can utilize growing databases of rare diseases to identify and prioritize protein coding variants that may impact kidney function but are anticipated to have more subtle phenotypes. We show that kidney tubular epithelial cells, in addition to podocytes, are impacted by the I1109V variant, suggesting a significant clinical impact beyond that predicted by modeling programs. Further, our results provide insight regarding how epithelial cell-based kidneys harboring variants of proteins affecting the cytoskeleton could develop normally but are at increased subsequent risk for cell injury, especially following exposure to stressors such as TNF-α or mechanical stress.

This study also reinforces the importance of utilizing complementary models to study the effects of variants of unknown significance in human disease, a critical step in the personalization of medicine. Here we relied on kidney organoids generated from iPSCs from an individual with disease, plus epithelial cell modeling with *X. laevis* embryos to reveal the impact of I1109V. Live cell imaging of *X. laevis* epithelial cells was key to demonstrating how the 1109V variant affects membrane morphology and fluctuation, actin cytoskeletal dynamics and barrier function, findings that would have been missed in a static system alone. Moreover, our established pipeline and platform could be used to study additional ANLN coding variants as well as other proteins associated with kidney disease and their variants.

First, we were not able to confirm the specificity of I1109V’s impact in kidney organoids by gene editing the human iPSCs to revert to WT ANLN, a time and resource intensive process. However, our complementary studies in *X. laevis* embryos employed ANLN KD and replacement with WT ANLN or the 1109V variant within the same clutches of embryos. Second, we did not directly test how 1109V affects ANLN’s ability to bind to lipids. In the future, this could be investigated using purified full-length ANLN or a PH domain fragment and carrying out binding assays with artificial membranes of various lipid compositions with a focus on PIP_2_-rich lipids. Third, we did not examine whether 1109V negatively affects tubular epithelial cell cytokinesis and successful tubular cell repopulation; future studies are warranted as this could have implications for recovery following acute tubular necrosis. Fourth, we did not explore whether I1109V is a restrictive variant given its single amino acid change and location within an alpha-helix of ANLN’s PH domain, which could impact 3D protein structure or interactions with other cellular components. In the future, this could be explored by introducing alternative residues at position 1109. Finally, we did not assess the impact of I1109V on epithelial cells of other organs such as the intestine. An intriguing speculative clinical scenario is that ANLN variants also affect intestinal barrier function and could result in uptake and systemic exposure to toxins which could also increase risk for CKD.

With increasing use of whole genome and whole exome sequencing to diagnose and characterize disease in the clinical realm, clinicians are accumulating information regarding coding variants in genes of interest with unknown significance (*51, 52*). Further information is often provided by predicting the potential impact of these variants on a protein’s function using available prediction tools (*53, 54*); however, experimental studies validating predictions are often lacking. Given the number of coding variants reported, pursuing studies identifying a molecular mechanism and impact of a specific variant using traditional research methods is impractical for patient care, requiring years of effort and significant expense. For variant impact assessment to be clinically impactful and timely, continued wide access to all research tools and model systems is critical, with the approach tailored to the disease and protein of interest. Therefore, creation of screening strategies utilizing combination of human cell-based organoids with model systems such as *X*. *laevis* embryos where mRNAs encoding multiple variants of unknown significance can be easily microinjected and phenotypes can be robustly and rapidly screened offers a pathway to expedite discovery. Furthermore, this study benefitted from connecting our findings with the novel ANLN I1109V variant in kidney organoids with significant previous work on ANLN’s cell biological functions in *X. laevis* embryos, demonstrating the critical need to support continued efforts to understand cell biology.

Potential specific therapies to mitigate I1109V’s effects on epithelial cell morphology and function were not explored in our study, in part due to the atypical nature of the scaffolding protein ANLN as a therapeutic target. Identifying effective treatments is especially challenging for atypical therapeutic targets. However, continued funding from NIH research programs such as the National Center for Advancing Translational Sciences’ focus on innovative strategies to treat undruggable targets within human disease could make these lofty goals achievable.

## MATERIALS AND METHODS

### Study Design

The objectives of this study were to identify Anillin variants in individuals with ProtKD and to characterize the cellular and molecular impact of the I1109V ANLN variant using kidney organoids and *X. laevis* embryos as model systems (Fig. 2D). We used WGS from the NEPTUNE cohort to identify *ANLN* variants and established correlations between *ANLN* mRNA expression and eGFR and UPCR in glomerular and tubulointerstitial tissue. A NEPTUNE participant-derived iPSC line with the heterogenous I1109V ANLN variant was used to generate kidney organoids. Single cell RNA-seq and immunohistochemistry were used to determine altered pathway expression, apoptosis and tubule cell morphology compared to control kidney organoids derived from a WT iPSC cell line. To investigate real-time epithelial cell junction and cytoskeletal alterations, *X. laevis* embryos were injected with an anti-sense morpholino, knocking down endogenous ANLN, and mRNAs encoding WT ANLN or the ANLN 1109V variant were microinjected into the embryos. Live cell imaging of *X. laevis* embryos allowed for quantification of junction waviness, transverse fluctuation, response to mechanical perturbation, and barrier function in the ANLN variant compared to WT. Detailed experimental designs, methodology, data analysis, and biological replicate numbers are described in the associated figure legends and Supplementary Materials and Methods.

### Statistical Analysis

Software used to present data and perform statistical tests are described in the Supplementary Materials and Methods. Experimental sample sizes and statistical tests used are noted in the associated figure legends and Supplementary Materials and Methods.

### List of Supplementary Materials

References (55–77)

Materials and Methods

Fig. S1 to S10

Movie S1 to Movie S3

Table S1 to S2

NEPTUNE Addendum

Data file S1

## Data Availability

Data produced in the present study are either available online at NCBI GEO repository or NephroSeq websites, contained in the supplemental materials, or available upon reasonable request to the authors pursuant to NIH, NEPTUNE Study and U-M data sharing requirements.

https://www.ncbi.nlm.nih.gov/geo/query/acc.cgi

https://nephroseq.org/resource/login.html

## ACKNOWLEDGEMENTS

We acknowledge all investigators and participants involved in the ERCB–Kröner-Fresenius Biopsy Bank (ERCB-KFB) and the NEPTUNE consortia. The Nephrotic Syndrome Study Network (NEPTUNE) is alumni of the Rare Diseases Clinical Research Network (RDCRN), which is funded by the National Institutes of Health (NIH) and led by the National Center for Advancing Translational Sciences (NCATS) through its Division of Rare Diseases Research Innovation (DRDRI). NEPTUNE has been funded under grant number U54DK083912 as a collaboration between NCATS and the National Institute of Diabetes and Digestive and Kidney Diseases (NIDDK). Additional funding and/or programmatic support is provided by the University of Michigan, NephCure Kidney International, Alport Syndrome Foundation and the Halpin Foundation. RDCRN active consortia and alumni are supported by the RDCRN Data Management and Coordinating Center (DMCC), funded by NCATS and the National Institute of Neurological Disorders and Stroke (NINDS) under U2CTR002818.

We are grateful to the NIH-funded National *Xenopus* Resource (RRID: SCR_013731) and Xenbase (RRID: SCR_003280) for public support of *Xenopus* research. The antibody to *X. laevis* ANLN was generously provided by Aaron Straight, Stanford University.

This project was supported by the University of Michigan Human Stem Cell Gene Editing Core Facility (RRID:SCR_026755), School of Medicine Advanced Genomics Core Facility (RRID:SCR_025788), BRCF Microscopy Core Facility (RRID:SCR_026722), the George M. O’Brien Michigan Kidney Translational Resource Center (RRID:SCR_015270) and Michigan Kidney Translational Medical Center (RRID:SCR_015903); the last two funded by NIH U54DK137314.

## Author contributions

Conceptualization: ALM, JLH

Data curation: DF, MF, CCB, MA, S. Wheeler, ML, FE

Formal analysis: ZC, HMJ, XML, CCB, JAS, SE, MA, S. Wheeler, FA, ML, RM, S. Weng

Methodology: ZC, HMJ, MF, CCB, MA, BG, FE, AM

Investigation: ZC, HMJ, MF, CCB, JAS, JES, MA, VV-W, FE, AM

Software: DF, S. Wheeler, FA

Supervision: MK, S. Weng, ALM, JLH

Validation: ZC, HMJ, MF, JAS

Visualization: ZC, HMJ, MF, XML, CCB, JAS, SE, MA, S. Wheeler, FA, RM

Resources: JES, BG, RM, MK

Funding acquisition: MK, S. Weng, ALM, JLH

Writing–original draft: ZC, HMJ, ALM, JLH

Writing–review & editing: ZC, HMJ, DF, MF, XML, CCB, JAS, JES, SE, MA, S. Wheeler, AM, VV-W, BG, FA, ML, FE, RM, MK, S. Weng, ALM, JLH

## Data, code, and materials availability

All data associated with this study are present in the paper or the Supplementary Materials. Next-generation sequencing data have been uploaded to the NCBI GEO repository: scRNAseq (GSE316404) and bulk RNAseq (GSE314915). Previously published data sets from the NCBI GEO repository were used for comparison as noted in the Supplementary Materials and Methods (GSE213972 and GSE230848). NEPTUNE RNA-seq and ERCB RNA-seq data are available at Nephroseq.org (*8, 68*). Cell cluster marker lists for scRNAseq data sets and lists of DEGs used for the analysis of scRNAseq and bulk RNAseq data are detailed in Data File S1. Figure illustrations generated using BioRender are publicly viewable at the following unique figure URLs: Fig. 2B (https://BioRender.com/duaoc2w), Fig. 2D (https://BioRender.com/2d4z5ir), Fig. 3B (https://BioRender.com/uwe4l1d), Fig. 3C (https://BioRender.com/sy0qv70), Fig. 8 (https://BioRender.com/yxdgg5p). Cell lines are available upon request under a material transfer agreement. All other materials used or generated in this study are commercially available or will be supplied upon reasonable request.

