## Supplemental Materials, Methods & Figures for "Anillin variant in proteinuric kidney disease drives tubular epithelial cell death, junctional instability, and barrier dysfunction"

**List of Supplementary Materials**
References (55–77)
Materials and Methods
Fig. S1 to S10
Movie S1 to Movie S3
Table S1 to S2
NEPTUNE Addendum
Data file S1

**SUPPLEMENTARY MATERIALS AND METHODS:**

**Whole Genome Sequencing in NEPTUNE**
Whole Genome Sequencing (WGS) was performed on 864 NEPTUNE participants. Reads were obtained using Illumina HiSeq 4000 System. Sequencing was performed to an average read depth of 30X. FASTQ files were evaluated for read quality using FastQC (version 0.11.9) and FastQScreen (version 0.15.1) (*55, 56*) and adapter trimming was done using FastP (*57*) (version 0.24.0). Reads were then aligned to the GRCh38 draft of the human genome using bwa-mem (*58*) (version 0.7.17). Joint variant calling and filtering was performed using GATK (*59*) (version 4.1.8.0) following recommended best practices as of (2024-07-24). Variants were then annotated using SnpEff (*59*) (version 5.0 build 2020-10-04).

**Variants in *ANLN***
Variants were extracted from the genomic coordinates of the *ANLN* gene (chr7:36389821-36453791 in GRChv38) flanked on either side by 1000bp. These were then filtered down to coding variants predicted to have high or moderate impact on the protein product. Listed gnomAD frequencies are current as of August 2025 (highest allele frequency from any individual genetic ancestry group). Nucleotide and amino acid locations based on the MANE Select transcript for ANLN (ENST00000265748.7/NM_018685.5).

**Monogenic Nephrotic Syndrome Variants**
The WGS from the seven NEPTUNE participants with the ANLN I1109V variant were also queried for variants in genes known to be associated with proteinuric kidney disease or genes in *ANLN*-associated Reactome (*60*) pathways (Tables S1-S2). Variants were extracted from a select list of genes. They were then further filtered using the following criteria: NEPTUNE allele frequency < 0.05, GERP (*61*) score > 3 and a predicted deleterious consequence according to either SIFT (*62*), MUTATION_Taster (*63*), or PolyphenHVAR (*64*).

**Transcriptome profiling**
In NEPTUNE, RNA sequencing (RNA-seq) was performed on manually microdissected kidney biopsy tissue that separated tubulointerstitial and glomerular compartments of the research core. Each compartment was profiled to obtain bulk glomerular and bulk tubulointerstitial RNAseq expression data profiles. Briefly, mRNA samples were prepared using the Illumina TruSeq mRNA Sample Prep v2 kit. Multiplex amplification was used to prepare cDNA with a paired-end read length of 100 bases using an Illumina HiSeq2000. RNAseq was performed by

the University of Michigan Advanced Genomics Core. Quality of the sequencing data was assessed using the FastQC. Read counts were extracted from the fastq files using HTSeq (version 0.11). Read counts were normalized by voom transformation (*65*). Data by biopsy proven diagnosis were plotted in GraphPad Prism v10. "Other" category in Fig. 1C-E includes individuals with IgA nephropathy, diabetic nephropathy, membranoproliferative glomerulonephritis, immune complex glomerulonephritis and those who presented clinically with nephrotic syndrome at enrollment; but without a pathology biopsy proven diagnosis recorded in the clinical data file at time of analysis.

Estimated glomerular filtration rate (eGFR) was calculated from serum creatinine measurements using CKiDU25 (*66*) and CKD-Epi equations (*67*) for participants below and above 25, respectively. Urine protein to creatinine (UPCR) ratios were centrally measured. eGFR and UPCR measurements closest to time of biopsy were log transformed and were used for correlation analyses (Pearson) against voom transformed ANLN expression in Spotfire (10.6.1).

Voom transformed ANLN expression was correlated (Pearson) with glomerular and tubulointerstitial TNF scores, representative of an active inflammatory state consistent with TNF activation. The TNF score, its components and methods for computing the TNF score have previously been described elsewhere (*8, 25*).

NEPTUNE RNA-seq and ERCB RNA-seq data are available at Nephroseq.org and through NCBI Gene Expression Omnibus (GSE219185, GSE197307) (*8, 68*).

**Organoid culture, treatment and sample preparation**
UM77-2 human (female) embryonic stem cells – ANLN WT (NIH approval #NIHhESC-14-0278, sourced from MStem Cell Laboratory) and NEPTUNE human (male) iPSC line 30B – ANLN I1109V (HUM00158219, HUM00120448, reprogrammed at the University of Michigan's Human Stem Cell & Gene Editing Core) were cultured in accordance with University of Michigan's Human Pluripotent Stem Cell Research Oversight Committee and NIH regulations. CNV assessment by Illumina bead array was performed on UM77-2 cells in 2022 at University of Michigan BRCF Advanced Genomics Core (RRID:SCR_025788) and confirmed normal karyotyping and sex. For 30B iPSCs, CNV assessment was performed by Illumina bead array (RRID:SCR_025788) and cell characterization testing was performed by WiCell Research Institute, both of which confirmed normal karyotyping and sex.

To confirm heterogenous ANLN variant at amino acid position 1109 (base pair 3325), the surrounding region was amplified using gDNA from UM77-2 and 30B iPSC cell lines and was submitted for Sanger sequencing (Azenta). Sequences were aligned to reference DNA using Benchling. Two primer sets were used: 1) forward: CCTAGCAAGAGTACATGGGGG, reverse: TCTCCAACCCCTGAAAGACG and 2) forward: TCCCTAGCAAGAGTACATGGG, reverse: TGCAATCAGTAAATCTGATGCTC.

Kidney organoids were generated using UM77-2 and 30B iPSCs as previously described (*25*) with the following alteration: 10 μM CHIR99021 used for organoid differentiation (30B:

Reprocell, #04000402 and UM77-2: StemCell Technologies, #72052). Organoids were cultured in RB media [Advanced RPMI (Gibco, #12633020) + Glutamax (Gibco, #35050061) + B27 Supplement (Gibco, #17504044) + Penicillin-Streptomycin (Gibco, #15140122)] from day 5 onward and all treatments were administered in RB.

TNF-α treatment: organoids were collected on day 24 or 25 following treatment with 5 ng/ml recombinant human TNF-α (R&D Systems, #10291TA050) reconstituted in DPBS (Gibco, #14190144; vehicle control) for 48 h (medium replenished every 24 h) prior to collection. Vehicle control (VC) for these experiments was 0.25-1.25% DPBS (v/v). Samples collected for scRNA-seq were collected on day 25 following 48h TNF-α treatment.

TNF-α + tacrolimus (FK506) treatment: organoids were collected on day 25 following treatment with 5 ng/ml recombinant human TNF-α reconstituted in DPBS and/or FK506 (MedChemExpress, #HY-13756) reconstituted in DMSO (Millipore Sigma, #D2650) for 48 h. Medium was replenished every 24 h prior to collection. VC for these experiments was 0.05% DMSO (v/v).

Latrunculin B treatment: organoids were collected on day 23 following treatment with 1 or 5 µM Latrunculin B (Tocris, #39-741) reconstituted in DMSO for 6 h prior to collection. VC for these experiments was 0.05% DMSO (v/v).

Jasplakinolide treatment: organoids were collected on day 24 or 25 following treatment with 0.5 or 5 µM jasplakinolide (Cayman Chemical, #11705) reconstituted in DMSO (vehicle control) for 1.5 h prior to collection. VC for these experiments was 0.175% DMSO (v/v).

**Organoid qRT-PCR**

Total RNA was collected from whole well organoid cultures as previously described (*25*). TaqMan Pre-Developed Assay Reagents (PDARs) used as follows: CXCL10, Hs00171042_m1; GAPDH, Hs03929097_g1 (ThermoFisher).

**Organoid immunofluorescence and microscopy**

Organoid spheroids were immunostained using methods previously described (*25*). Samples were imaged using a Leica DM IRB microscope with Olympus DP70 digital camera, a Nikon Ti2 Widefield, or a Zeiss LSM 980 Airyscan 2 (Michigan Medicine Microscopy Core).

Primary antibodies, lectins and molecular probes used: Anillin (ThermoFisher Scientific, PA5-18199, 1:50, RRID:AB_10980141), Cleaved Caspase 3 (Cell Signaling Technology, 9661T, 1:400, RRID:AB_2341188), Collagen 1 (Abcam, clone EPR7785, ab138492, 1:2000, RRID:AB_2861258), Cytokeratin, pan (Millipore Sigma, C2562, 1:500, RRID:AB_476839), E-cadherin (BD Biosciences, clone 36/E-Cadherin, 610181, 1:200, RRID:AB_397580), Fibronectin 1 (Millipore Sigma, F3648, 1:400, RRID:AB_476976), K-cadherin (R&D Systems, clone # 427909, MAB2715, 1:500, RRID:AB_907139), MEIS1/2/3 (Active Motif, clone 9.2.7, 39795, 1:300, RRID:AB_2750570), N-cadherin (R&D Systems, AF6426, 1:1000, RRID:AB_10718850), Nephrin (R&D Systems, AF4269, 1:500, RRID:AB_2154851), Phalloidin (Invitrogen, O7466,

1:1000), Podocalyxin (R&D Systems, AF1658, 1:1000, RRID:AB_354920), Wheat germ agglutinin (Vector Laboratories, FL-1021, 1:1500, RRID:AB_2336866), Zonula occludens 1 [ZO-1] (Invitrogen, clone ZO1-1A12, 33-9100, 1:100, RRID:AB_87181).

Secondary antibodies used (ThermoFisher Scientific, 1:500): Alexa Fluor 488 - Donkey anti-Mouse (A-21202, RRID:AB_141607), Donkey anti-Rabbit (A-21206, RRID:AB_2535792), Donkey anti-Goat (A-11055, RRID:AB_2534102), Donkey anti-Sheep (A-11015, RRID:AB_2534082); Alexa Fluor 594 - Donkey anti-Mouse (A-21203, RRID:AB_2535789), Donkey anti-Rabbit (A-21207, RRID:AB_141637), Donkey anti-Goat (A-11058, RRID:AB_2534105), Donkey anti-Sheep (A-11016, RRID:AB_2534083); Alexa Fluor 647 - Donkey anti-Mouse (A-31571, RRID:AB_162542), Donkey anti-Rabbit (A-31573, RRID:AB_2536183), Donkey anti-Goat (A-21447, RRID:AB_2535864).

**Organoid microscopy quantification**

FIJI (*69*) was used to analyze Nikon (.nd2) files and Zeiss (.czi) Airyscan-processed files. Sum images were generated by using the Z project tool to sum 15 slices. Images were captured using the same microscopy settings within each experiment. Micrographs presented in the figures were contrast adjusted to the same levels within the individual figure part presented for comparison.

QuPath v0.6.0 or older (*70*) was used to analyze epifluorescence images. Pan-cytokeratin IF staining, which recognizes all tubule structures, was used to quantify tubular area in QuPath. The closed polygon tool was used to outline the exterior perimeter of tubule structures, followed by outlining the interior lumen perimeter. Using these two paired area measurements ("tubular area" and "lumen"), the percentage of the tubular area taken up by the lumen versus the tubular cells could be calculated. To measure areas positive for a specific stain, the brush tool was used to annotate the area of interest. A thresholder was created to delineate positive and negative regions within annotations. Area measurements of positive regions were used to calculate the stain-positive percentage of each annotation. Due to fluorescence intensity variations between image collection days, data presented from multiple experiments is displayed as fold-change difference between VC and TNF-α treatment for independent experiments. To quantify the number of apoptotic tubular cells, tubules were annotated using the above method, cells staining distinctly for cleaved caspase 3 within each tubular cell annotation were manually counted, then divided by the tubular cell area to generate the number of apoptotic cells per tubule area. GraphPad Prism v10.0 or higher was used to present IF quantification data and perform statistical tests. When comparing two groups, a two-tailed t-test or Welch's t-test was used. A one-way ANOVA with Šídák's multiple comparison test was used when comparing more than two groups and results from all comparisons made using this test were reported. Statistical analyses performed are described in the associated figure legends.

**Organoid scRNA-seq and bioinformatic analysis**

24- or 25-day-old organoids were dissected from 24 well plate kidney organoid cultures, washed with ice-cold DPBS and dissociated with cold active protease (Sigma, P5380). Following dissociation, the single cell suspensions were resuspended in DMEM/F12 (Gibco, #11320033) +

0.1% BSA (Gibco, #15260037) and then submitted to the Advanced Genomics Core at the University of Michigan for library preparation and sequencing on a 10x Genomics Chromium System. Organoid scRNA-seq data was processed as previously described (*25*). Data available at NCBI GEO accession number GSE316404. 3549-JES-1 from GSE213972 and 3748-JES-1 from GSE230848 were used for comparison to 6837-MF-1. Cell cluster markers list for ANLN WT vs ANLN I1109V kidney organoids is provided in Data File S1.1. Cell cluster markers list for ANLN I1109V kidney organoids VC vs. 48h TNF-α treatment (5 ng/mL) is provided in Data File S1.4.

Differential expression (group 1: 6837-MF-1 vs. group 2: JES3549, JES3748) was performed independently in tubular epithelial cells and podocytes. Within each cell type, differential expression was carried out comparing group 1 to group 2 samples. Single cell differential expression was carried out using the FindMarkers function in the Seurat R package (version 5.3.0). Only genes expressed in at least 10% of the cells in either population were analyzed (min.pct = 0.1 flag). See Data File S1.2 for list of 4,109 genes and top 500 DEGs in tubular epithelial cells. See Data File S1.3 for list of 3,587 genes and top 500 DEGs in podocytes.

Literature-based gene networks from the genes differentially regulated (as defined by our filter criteria; see Data File S1.2 and S1.3) were generated using the Genomatix Pathway System (GePS) software installed on a local server (Genomatix Genome Analyzer (GGA), version v3.150510). In these networks, the 50 best connected genes co-cited in PubMed abstracts in the same sentence linked to a function word (most relevant genes/interactions) are shown.

**_Xenopus laevis_ embryos**

Adult, female *X. laevis* (African Clawed Frog; *Xenopus* 1 or National *Xenopus* Resource (NXR)) were injected with human chorionic gonadotrophin (HCG; MP Biomedicals, #198591) to induce hyper-ovulation. Eggs were then collected from the frogs the next day and fertilized *in vitro* using excised testes from male frogs. Fertilized embryos were maintained in 0.1X MMR (10 mM NaCl, 0.2 mM KCl, 0.2 mM $CaCl_2$, 0.1 mM $MgSO_4$ and 0.5 mM HEPES, pH ≥ 7.4). Thirty min following fertilization, embryos were de-jellied using 2% cysteine in 1X MMR until the jelly coat was removed and then transferred to 0.1X MMR and allowed to develop to the 2- or 4- cell stage for microinjection. All experiments conducting *X. laevis* frogs and embryos were compliant with the University of Michigan Institutional Animal Care and Use Committee (IACUC) and the U.S. Department of Health and Human Services Guide for the Care and Use of Laboratory Animals.

**Microinjection of _X. laevis_ embryos**

Microinjection experiments were carried out using a BTX pico-injector microinjection apparatus (Harvard Apparatus). Glass capillary needles were manually calibrated to eject 5 nL of liquid per one-second injection. Embryos allowed to develop to the 2-cell stage were injected for one second twice per blastomere with morpholino and/or mRNAs for a total injection volume of 20 nL. Embryos allowed to develop to the 4-cell stage were injected with morpholino and/or mRNAs for one second, once per blastomere for a total injection volume of 20 nL.

***Anln* anti-sense morpholino**

A custom morpholino oligomer (Morpholino sequence: 5' TGGCTAGTAACTCGATCCTCAGACT 3') that targets the 5'UTR of *anln* mRNA transcripts was ordered from GeneTools as previously described (*19-21*).

**Construction of the *X. laevis* ANLN 1109V variant DNA construct**

The Q5 Site-Directed Mutagenesis Kit (NEB, #E0554S) was used to make the desired nucleotide mutations in *X. laevis* WT ANLN sequence to generate the L1109V mutation. The NEB base changer website was used to design primers for use with the mutagenesis kit (Forward 5':TCTTGTGGATGTGCGAATGTGGC ; Reverse 3': ACTTGATTCAGCTTTTGCATC). pCS2+ vector-based untagged ANLN L1109V, ANLN L1109V-mNeon and ANLN L1109V-Halo constructs were made. Following transformation into DH5α cells, colonies were selected and mini-prepped, then sent for full-plasmid sequencing (Plasmidsaurus) to confirm the desired mutation was successfully made as we all as the absence of off-target mutations.

***in vitro* transcription of mRNA**

The ANLN plasmids were linearized using Kpn1, *in vitro* transcription was carried out using the mMessage mMachine SP6 Transcription Kit (Fisher, #AM1340) and the resulting mRNA was purified using the RNeasy Mini Kit (Qiagen, #74104). Purified mRNAs were stored at -80ºC at a 5X concentration until use, where they were diluted to a 1X injection concentration in combination with additional tagged proteins of interest, fluorescent probes, or morpholino oligos as indicated below. All other DNA constructs were previously described constructs from the Miller lab strain collection: mCherry-farnesyl (*19*), membrane-BFP (*71*), BFP-ZO-1 (*42*), Halo-ZO-1 (*72*), LifeAct-mRFP (*71*).

**Bulk RNAseq of *X. laevis* embryo**

<u>RNA sample collection and processing:</u> *X. laevis* embryos at the 2- or 4- cell stage were injected with either ANLN morpholino (KD) or water control (WT). Following injection, embryos were allowed to develop overnight at 15°C. Total RNA was collected from 5 embryos per condition, in duplicate and RNA was extracted using Direct-zol RNA miniprep kit. Library prep and next-generation sequencing of total RNA on a NovaSeq6000 were carried out in the Advanced Genomics Core at the University of Michigan.

<u>Read alignment, quality control and gene-level quantification:</u> Raw FASTQ reads were aligned to the *Xenopus laevis* reference genome (GCF_017654675.1; genome assembly v10.1; Xenbase v10.1) using a standard RNA-seq alignment workflow. Quality control (QC) was performed at the read, alignment and dataset levels following a previously described RNA-seq QC pipeline (*73*). Read-level QC evaluated sequencing quality metrics (e.g., per-base quality scores, GC content and adapter contamination). Alignment-level QC assessed mapping performance and read distribution across genomic features and dataset-level QC evaluated global expression patterns to identify potential technical outliers prior to downstream analyses. Gene-level quantification was performed with HTSeq by assigning reads to annotated *X. laevis* genes using the corresponding genome annotation, yielding a raw gene-by-sample count matrix.

Principal component analysis (PCA): PCA was performed to visualize global transcriptional variation across samples using normalized gene-level expression values. The expression matrix (genes × samples) was transposed (samples × genes), centered and analyzed with prcomp() in R. The first two principal components (PC1 and PC2) were used for visualization and the percent variance explained was computed from the eigenvalues and displayed on axis labels.

Normalization and differential expression analysis: Raw counts were normalized using the voom transformation to model the mean–variance relationship and compute precision weights for linear modeling. Differential expression between knockdown (KD) and wild-type (WT) samples was tested using limma (*74*). *p*-values were adjusted for multiple testing with the Benjamini–Hochberg false discovery rate (FDR) and differentially expressed genes (DEGs) were defined as FDR < 0.05. At this threshold, 1550 genes were differentially expressed. See Data File S1.5.

Visualization of ANLN expression: Voom-normalized expression values for *X. laevis anln*.L were extracted and samples were grouped as WT or KD based on experimental metadata. Group-wise expression was visualized using ggplot2 boxplots (median, interquartile range and whiskers extending to 1.5×IQR). The y-axis represents voom-normalized expression ($\log_2$ counts per million with precision weighting).

Reactome pathway enrichment analysis: Reactome pathway enrichment was performed using ReactomePA with pathway annotations provided by Reactome.db (v1.89.0). Because Reactome annotations are not available for *X. laevis*, DEGs were mapped to human orthologs using the Xenbase GeneGenomeHumanOrtho pivot table. Each *X. laevis* symbol was first matched directly (retaining the ".L/.S" homeolog suffix); if no match was found, the suffix was removed and the base symbol was matched. 1030 valid human genes were recovered from the initial 1550 DEG *X. laevis* genes. Human Entrez Gene identifiers were obtained for mapped orthologs using org.Hs.eg.db when needed. Rank-based gene set enrichment analysis was then performed with gsePathway() (*75*), ranking genes by the *Xenopus*-derived $\log_2$ FC. Statistical significance was assessed using Benjamini–Hochberg FDR correction and pathways with *p*-value<0.1 were considered significant.
Bulk RNAseq data available at NCBI GEO accession number GSE314915.

**Western blot of *X. laevis* embryos**
Embryos were injected with either water (control) or 2.5 mM of the *Anln* morpholino at the 2- or 4-cell stage and allowed to develop overnight at 15 ºC. Embryos were then sorted to remove any damaged or dead embryos, and number-matched between groups. Embryos were then washed once in 200 µL of PHEME lysis buffer (60 mM K-PIPES, 25 mM HEPES, 10 mM EGTA, 2 mM $MgCl_2$, pH 7.0) with 0.5% NP40, 3% protease inhibitor cocktail (Thermo Scientific #PI78410), and 3% phosphatase inhibitor cocktail (Thermo Scientific, # PI78420) and then lysed in 5 µL/embryo of the same solution used for washing and homogenized using a pestle. Lysed embryos were transferred to an Ultra-Clear tube (Beckman Coulter, #344718) and centrifuged at 14,000 rpm for 5 minutes at 4 °C to separate the lysate. The cytoplasmic layer was isolated by puncturing the tube with a PrecisionGlide 27 G needle (BD, #305109) and drawing the lysate

into a 1 mL syringe, and the volume of lysate was equalized between samples. ½ volume of 6X SDS was then added to the cytoplasmic lysate, and the samples were boiled for 10 minutes. Samples were run on 4–15% Mini-PROTEAN® TGX™ Precast Protein Gels (Bio-Rad, #4561086) and wet transferred onto PVDF membranes (MilliporeSigma, #IPVH08100) overnight at 4 °C at 15V. Membranes were blocked in 5% milk in TBST for one hour at room temperature with shaking, then incubated with anti-Anillin antibody (1:1,000, gifted from Aaron Straight, Stanford University (*76*)) and anti-tubulin antibody (1:15,000, Sigma, #T9026) overnight at 4 °C. Membranes were then washed 3 times for 15 minutes each in TBST, and incubated with horseradish peroxidase secondary antibodies: anti-mouse IgG-HRP (1:5,000, Promega, #W4011), anti-rabbit IgG-HRP (1:5,000, Promega, #W4021) for 2 hours. Membranes were washed 3 times for 15 minutes each in TBST, then washed for 5 minutes in PBS, and then developed immediately prior to imaging using SuperSignal West Pico Chemiluminescent Substrate (Fisher, # PI-34077).

**Confocal imaging of *X. laevis* embryos**

Microscopes: Embryos were imaged using an Olympus FluoView 1000 or FluoView 3000 scanning confocal microscope, with a 60X supercorrected Plan ApoN60XOSC objective (NA = 1.4, working distance = 0.12 mm).

Fixed immunofluorescence imaging: Embryos were co-injected at the 2- or 4-cell stage with 2.5 mM of the *anln* morpholino and 200 pg of either WT or L1109V *anln* mRNA (untagged) as well as 50 pg of mCherry-farnesyl mRNA as a lineage tracer so that we could identify cells that had received the morpholino and rescue construct. Embryos were allowed to develop overnight at 15ºC and then fixed for 2 h at room temperature in 2% trichloroacetic acid (TCA) and PBS. After three washes in 1X PBS, embryos were bisected into animal and vegetal hemispheres; the vegetal hemisphere was discarded. The animal hemisphere was permeabilized in 1% Triton X-100 in PBS for 20 minutes, 0.1% Triton X-100 in PBS for 20 minutes, blocked in 5% FBS in PBST for 1 h at room temperature without shaking and then incubated overnight at 4ºC in primary antibodies: 1:200 α-mCherry (mouse, Abcam, ab125096) and 1:200 α-ZO-1 (rabbit, Invitrogen, 61-7300). Embryos were then washed in 0.1% PBST for 5 min, 15 min and 2 h and blocked overnight at 4ºC. After a quick wash, embryos were incubated at 4ºC with appropriate secondary antibodies 1:200 (goat anti-mouse Alexa Fluor™ 568, Invitrogen, A-11004; goat anti-rabbit Alexa Fluor™ 647, Invitrogen, A-21244) for 6 h, then washed in 0.1% PBST for 5, 10 and 15 min before storing in blocking solution overnight at 4ºC. On the final day, embryos were washed once in 0.1% PBSN (NP40, PBS) then incubated with 1:1000 DAPI in TBSN, then washed every 10 min for 1 h with 0.1% TBSN (NP40, TBS). Finally, embryos were stored in 1X PBS. When imaging, fixed embryos were placed on a glass slide and a coverslip was attached using vacuum grease and then sealed with clear nail polish. Samples were imaged by acquiring 0.5 µm z-sections through all detectable signal from apical to basal.

Live imaging: Embryos were co-injected at the 2- or 4-cell stage with 2.5 mM *anln* morpholino and 200 pg of either WT *anln*-mNeon or *anln* L1109V-mNeon mRNA, 70 pg BFP-ZO-1 mRNA (tight junction marker) mRNA and 45 pg of LifeAct-mRFP (F-actin probe) mRNA. Following injections, embryos were kept at 15ºC and allowed to develop overnight. At gastrula stage, embryos were mounted in a metal slide (~0.9 mm thick with a hole in the center) such that the embryos were held in place between two coverslips attached to the slide with vacuum grease and live imaged.

Extracellular ATP treatment to increase actomyosin-mediated tension: Embryos were co-injected at the 2- or 4-cell stage with 2.5 mM *anln* morpholino and 200 pg of either WT *anln*-mNeon or *anln* L1109V-mNeon mRNA, 20 pg membrane-BFP, 20 pg LifeAct-mRFP and 150 pg of Halo-ZO-1 mRNA and 5 µM of Janelia Fluor® 646 HaloTag® Ligand (Promega, #GA1120). Embryos were allowed to develop overnight at 15ºC and then live-imaged as previously described, with the following modification: when mounted on metal slides, the top coverslip only covered half of the embryo, leaving an opening to add ATP solution. ATP solution was prepared by dissolving ATP (Sigma, A6419-1G) in 0.1X MMR at a 500 µm concentration. Live movies were acquired using the following parameters: 512x512 area, scan speed of 4 µs/pixel, zoom 2.0, step size 0.5 µm, imaging 6 of the most apical optical z-slices of the embryo. Videos were acquired for 8 frames and then 100 uL of ATP solution as pipetted into the open side of the slide. The video was acquired for a total of 60 frames (time interval between frames = 20 sec).

Zinc-based Ultrasensitive Microscopic Barrier Assay (ZnUMBA): Embryos were co-injected at the 2- or 4-cell stage with 2.5 mM ANLN morpholino and 200 pg of either WT *anln*-Halo or *anln* L1109V-Halo mRNA and 5 µM of Janelia Fluor® 646 HaloTag® Ligand (Promega, #GA1120), 70 pg BFP-ZO-1 mRNA and 45 pg of LifeAct-mRFP mRNA. Following injections, embryos were kept at 15ºC and allowed to develop overnight. The following day, embryos were injected in the blastocoel with 10 nL of 1 mM FluoZin3 (ThermoFisher Scientific, F24194), 100 µM $CaCl_2$ and 100 µM EDTA and allowed to recover for at least 5 min. At gastrula stage, embryos were mounted in 1 mM $ZnCl_2$ in their normal media (0.1xMMR) and live imaged with the following parameters: 512x512 area, scan speed of 4 µs/pixel, zoom 2.0, step size 0.5 µm, imaging 8 of the most apical optical z-slices of the embryo.

Quantification: Images were captured using the same microscopy settings within each experiment. Images presented in the figures were contrast adjusted to the same levels within the individual figure parts presented for comparison.

Randomization and blinding of data: Files were randomized and blinded for analysis of junction linearity, F-actin flares and FluoZin3 intensity using the RandomNames.bat script (https://www.howtogeek.com/57661/stupid-geek-tricks-randomly-rename-every-file-in-a-directory/). Blinded files were analyzed and then sorted into the correct experimental category following quantification using the true file identity translation.

Junction separation: Junction separation was quantified using TCA fixed images. In FIJI, a gray LUT was applied to the ZO-1 channel and all junctions in a field-of-view were counted with the multi-point tool. The number of separated junctions was then counted and divided by the total number of junctions in a field-of-view to give percent separation for an embryo. Separated junctions were defined by two distinct pools of ZO-1, separated by a negative space of no ZO-1 signal.

Junction linearity: Junction linearity was quantified using Junction Mapper (*77*). Using live-imaging movies, the first frame was isolated and made into a single maximum projection image. Image channels were pseudo colored green (ZO-1) and red (F-actin), merged, exported as a TIF file, and then files were randomized and blinded. The TIF image was loaded into Junction Mapper and put through the software's adaptive thresholds [C Value = 10, filter size = 99x99] to produce a binary image. Small, cytoplasmic, noise pixels were removed using the 'Remove Small Objects' function. The image was then skeletonized and overlaid on top of the original image to identify any gaps or aberrant lines within the skeletonized junctions, which could be added or removed manually. After finalizing the junction skeleton, cells were selected randomly across the field of view for measurement, prioritizing junctions that displayed abnormal morphology (e.g., bulging or wavy junctions) and excluding cells undergoing cytokinesis. Six cells were selected per embryo. To avoid quantifying junctions twice, the selected cells did not share bicellular junctions. The software generated a junction outline for each cell, and the vertices were manually added for each cell. The junction linearity index (ratio between the true length of the junction and the straight distance between the vertices of the same junction) was calculated and exported as an excel sheet. All analysis was performed in R, and graphs were produced using the package ggplot2.

Transverse fluctuation: We employed our recently published TFlux analysis pipeline (*33*) to quantify transverse movement along cell-cell junctions in ANLN KD+WT and ANLN KD+1109V embryos. Junctions were labeled with BFP-ZO-1 and live imaged at a spatial resolution of 0.441 μm/pixel and a temporal resolution of a ~20 sec/frame. This analysis included a broad range of junctions across the field of view, but those involved in cytokinesis were excluded due to the dynamic remodeling of ZO-1 near the cleavage furrows.
We used ilastik® Pixel Classification and ilastik® Carving to reconstruct the 3D (xyt) meshes of junctions from time-lapse movies. A customized MATLAB script was then used to quantify their transverse movement over time. Briefly, for each time point, we defined a baseline junction position by performing a spatial moving average over 2 μm and a subsequent temporal moving average over a 40 sec window. The transverse movement at each time point was measured as the distance between each pixel on the original junction and the corresponding temporally smoothed baseline (Fig. 5G).
Kymographs were generated to visualize the spatiotemporal patterns of the transverse movement (Fig. 5H-J). Distributions of absolute transverse movement were binned and normalized for per junction to generate heatmaps (Fig. 5K) and their pooled distribution was also calculated (Fig. 5L and Fig. S9A), showing a long-tailed increase in movement in mutant-rescued junctions. Cumulative probability plots, summarizing the probability up to a given magnitude along the x-axis, again revealed a clear rightward shift (Fig. S9B-C). Statistical significance was assessed using the Kolmogorov–Smirnov test on pooled data.

F-actin kymographs: Kymographs were created in FIJI. A three pixel-wide segmented line was traced around the junctions of a cell, then added to the ROI manager. The line was evaluated every few frames and manually repositioned when necessary to ensure continued overlap with the junctions during cell shape changes. The FIJI plugins ‘Timelapse’ and ‘Interpolate ROIs’ were used to generate ROI lines for all subsequent frames and add them to the ROI manager. Kymographs were then generated from the ROIs using ‘Analyze’, and the ‘Multi Kymograph’ plugin. Each line of the kymograph is the average of the three pixels from the ROI.

F-actin flare frequency: Files were randomized and blinded. F-actin flares were counted as an increase in intensity resulting in a color change within the Fire LUT (increase of at least ~45% from baseline) and lasted for at least 2 frames. Areas of stable increased intensity (longer than 6 frames) were not counted. Time interval between frames = 20 sec.

Junctional FluoZin3 intensity: The first frame (time = 0 min) and last frame (time = 30 min) of each video was used to create two maximum intensity projections (8 z-slices). Cell-cell junctions were manually traced using the FIJI brush tool (circular ROI 1.32 μm (3 pixels) wide) and measured to determine the total intensity of FluoZin3 signal at junctions at the start and end of the video. All videos were acquired using the same laser power for the FluoZin3 channel. One embryo was removed due to drift during image acquisition.

Statistical tests: GraphPad Prism v10.2 was used to plot data and perform statistical tests. Differences between control, ANLN KD, ANLN KD+WT and ANLN KD+1109V embryos were generally determined using an unpaired Student’s T-test. For junction separation: a one-way ANOVA was used to determine if there was a difference between control, ANLN KD, ANLN KD+WT and ANLN KD+1109V, while pairwise comparisons were made using an unpaired Student’s T-test. For junction linearity: the distribution of both ANLN KD+WT and ANLN KD+1109V were identified as non-parametric and were therefore compared using a Mann Whitney U test.

**Data presentation**

BioRender was used to generate scientific illustrations (in part [illustrations only]: Fig. 2B and 2D; Fig. 3B-C; in total: Fig. 8). Unique BioRender figure URLs are listed in the acknowledgements. Adobe Illustrator was also used to generate scientific illustrations (Fig. 4-7, S6, S8-10).

**SUPPLEMENTARY FIGURES:**

**Figure S1. *ANLN* in proteinuric kidney disease (protKD)**

**(A)** Two unique non-coding variants identified in five NEPTUNE study participants, predicted to highly impact ANLN protein function.

**(B)** WGS from the seven individuals with an I1109V *ANLN* variant were searched for mutations in other genes potentially related to protKD. A list of known monogenic contributors to protKD and genes in three Reactome pathways related to ANLN function were used for this search (Tables S1-S2). The red star and box indicate individual from which the iPSC line used for organoid generation in this study was derived.

**(C-D)** Scatter plots showing TNF score and *ANLN* mRNA abundance correlation in glomerular tissue (C) and tubulointerstitial tissue (D) from NEPTUNE study participants diagnosed with FSGS or MCD. Simple linear regression used.
**(E)** Bar graphs showing glomerular expression of *ANLN* mRNA in participants of the European Renal cDNA Bank (ERCB) with MCD compared to healthy living transplant donors.
**(F)** Bar graphs showing tubulointerstitial expression of *ANLN* mRNA in ERCB participants with MCD and FSGS compared to healthy living transplant donors. (E-F) Bulk RNAseq data from NephroSeq was used for these analyses. Unpaired t-tests were performed comparing healthy living donor to protKD group.

**Figure S2. I1109V kidney organoid characterization**
**(A)** Immunofluorescence (IF) imaging of sectioned kidney organoids generated from I1109V iPSCs produce epithelialized tubular cells (E-/N-/K-cadherin), podocytes (podocalyxin, nephrin) and stromal cells (MEIS1/2/3). Individual channels from Fig. 2E shown here in grayscale.
**(B-C)** IF imaging of sectioned untreated WT and I1109V kidney organoids. Tubule structures are stained with Pan-CK (red) and areas of fibrosis with (B) collagen 1 (COL1, green) or (C) fibronectin 1 (FN1, green). The percentage of total organoid area staining positive for fibrotic marker was quantified using QuPath. Points shown represent individual quantified organoids (n = 11-14) from one experimental replicate. Box plots depict median, $25^{th}$ and $75^{th}$ percentiles and min/max. *p*-values generated using two-tailed t-test. ns: $p \geq 0.05$.
**(D)** Dot plot showing expression of key cell type genes in single cell transcriptomes of D25 untreated I1109V and D24 vehicle control (PBS or DMSO) WT organoids. Data was visualized using CZ CELLxGENE.
**(E)** Literature-based network analysis from the top 500 differentially regulated genes (FDR <0.01, Abs $Log_2$ fold-change ≥1.0) in I1109V variant podocytes compared to wild-type. GePS literature-based network represents the top 50 best connections.

**Figure S3. Characterizing TNF-α treatment of I1109V kidney organoids**
**(A)** qRT-PCR for pro-inflammatory cytokine CXCL10 in WT and I1109V kidney organoids after TNF-α treatment. Data are means +/- SEM (n = 4). *p*-values generated using Welch's t-test. *$p \leq 0.05$, **$p \leq 0.01$.
**(B)** Quantification of tubular cell area in I1109V kidney organoids staining positively for ANLN following TNF-α treatment. Data plotted as fold-change increase in tubular cell area staining positively for ANLN (relative tubular ANLN+ area). Data are means +/- SEM *(*n = 3). *p*-values generated using Welch's t-test. ns: $p \geq 0.05$.
**(C)** IF imaging of sectioned I1109V kidney organoids comparing vehicle control and TNF-α treatment. Tight junctions in tubular cell areas are staining with ZO-1 (red) and F-actin is visualized using phalloidin (green). Dashed boxes highlight enlarged areas displayed to the right. Images are sum Z-projections of 1.95 µm. Cyan arrowheads on TNF-α-treated tubule micrograph (bottom) indicate cells with concentrated F-actin staining around their periphery
**(D-E)** Lumen area as a percentage of total tubular area quantified in I1109V kidney organoids. Proximal (N-cadherin+) and distal (E-cadherin+) tubules were compared between vehicle control (VC) and TNF-α-treated organoids. "All" group is comprised of both proximal and distal tubules. Mean data presented in (D). Data points in (E) represent individual quantified tubules (n

≥ 48 tubules ["All"] for VC and TNF-α) from two experimental replicates. Box plots depict median, 25th and 75th percentiles and min/max. *p*-values generated using two-tailed t-tests.

**Figure S4. Response of I1109V kidney organoids to TNF-α stress**

**(A)** Uniform Manifold Approximation and Projection (UMAP) representation of single cell transcriptomes of D25 untreated and TNF-α-treated I1109V kidney organoids distinguished 14 cell type clusters.

**(B)** Dot plot showing expression of key cell type marker genes in single cell transcriptomes of D25 I1109V untreated and TNF-α-treated kidney organoids verify cell type clusters. (A, B) Data visualized using CZ CELLxGENE.

**(C)** Method of quantifying areas of apoptotic podocytes in kidney organoids using IF images.

**(D)** IF imaging of WT and I1109V kidney organoids treated with TNF-α. Podocyte clusters are stained with podocalyxin (PODXL, red) and apoptotic cells with cC3 (green).

**(E)** Quantification of apoptotic podocytes per podocyte area based on IF staining (representative images show in (D)). Data are displayed as means +/- SEM (n = 2). *p*-values generated using one-way ANOVA with Šídák's test. All comparisons made are shown. ns: $p \geq 0.05$.

**(F)** IF imaging of sectioned WT kidney organoids treated with TNF-α and/or FK506. Tubule structures are stained with Pan-CK (red) and apoptotic cells with cC3 (green).

**(G)** Quantification of apoptotic cells per tubule area based on IF staining (representative images in (F)). Data are displayed as means +/- SEM (n = 3). *p*-values generated using one-way ANOVA with Šídák's test. All comparisons made are shown. ns: $p \geq 0.05$.

**Figure S5. Actin perturbation and stabilization in I1109V kidney organoids**

**(A)** IF imaging of sectioned WT and I1109V kidney organoids treated with LatB (1 μM or 5 μM, 6h). Tubule structures are stained with Pan-CK (red), F-actin is visualized using phalloidin (green) and ANLN is stained in magenta.

**(B)** IF imaging of sectioned I1109V kidney organoids treated with jasplakinolide (JAS; 1 μM or 5 μM, 1.5h). Tubule structures are stained with Pan-CK (red) and apoptotic cells with cC3 (green).

**(C)** Quantification of apoptotic cells per tubule area based on IF staining (representative images in (B)). Data are displayed as means +/- SEM (n = 2). *p*-values generated using one-way ANOVA with Šídák's test. All comparisons made are shown. ns: $p \geq 0.05$.

**Figure S6. The ANLN PH domain is well conserved between humans and *X. laevis* and validation of ANLN knockdown in *X. laevis embryos.***

**(A)** Human ANLN protein domain diagram. The location of the ANLN variant characterized in this paper, I1109V, is indicated with a red star. Myosin II binding domain (Myo), Rho binding domain (RBD) and pleckstrin homology domain (PH).

**(B)** Sequence alignment of human and *X. laevis* ANLN PH domains. Conserved residues are shown in dark orange, similar residues (by charge or polarity) are shown in light orange. Non-conserved residues are shown in gray. Position 1109, isoleucine in humans and leucine in *X. laevis*, is shown in yellow and marked with a red star.

**(C)** Table showing the percentage of amino acid identity and similarity between human and *X. laevis* for full length ANLN (left) or the ANLN PH domain (right). Sequences obtained from UniProt: *Homo sapiens*, Q9NQW6; *X. laevis*, Q801E2.
**(D)** Schematic depicting the morpholino oligo (MO) target sequence within the 5' UTR of *X. laevis anln*. The MO binds to the 5' UTR of *X. laevis anln* mRNA, blocking translation.
**(E)** Western blot showing ANLN protein levels in control (water-injected), *anln* KD (2.5 mM *anln* MO) and *anln* over-expression (OE, 200 pg mRNA injected) embryos. ANLN intensity was normalized to alpha tubulin intensity. 30 embryos were injected per group and the bulk embryo lysate was collected from 15 embryos, while the other 15 embryos were used for RNAseq analysis (Fig. S7).

**Figure S7. Bulk RNAseq of WT and ANLN KD *X. laevis* embryos**
**(A)** Principal component analysis of bulk RNAseq data from WT or ANLN KD *X. laevis* embryos.
**(B)** *anln* mRNA expression in WT or ANLN KD *X. laevis* embryos.
**(C)** Differentially regulated Reactome pathways in ANLN KD compared to WT after mapping RNAseq reads from *X. laevis* embryos onto human orthologs.

**Figure S8. ZO-1 immunofluorescence and examples of junction separation in *X. laevis* embryos.**
**(A)** IF imaging of ZO-1 in control, ANLN KD, ANLN KD+WT and ANLN KD+1109V embryos. Dashed boxes highlight reduced ZO-1 intensity in ANLN KD and junction separation in ANLN KD+1109V.
**(B)** IF imaging of ZO-1 showing representative examples of junction separation in an ANLN KD+1109V embryo. Areas of junction separation (yellow arrowheads) are present throughout the field of view. Areas that were not counted as junction separation (green dashed boxes) show a single line of ZO-1; in contrast, areas that were counted as junction separation (blue dashed boxes) show two distinct lines of ZO-1 signal.

**Figure S9. Transverse junction movement is increased in *X. laevis* embryos expressing ANLN 1109V.**
**(A)** Bar plots highlighting the long tail region of the same distribution shown in Fig. 5L.
**(B and C)** Cumulative probability plots of absolute transverse movement between 0-0.6 μm (B) and 0.6-2 μm (C). Junctions in ANLN 1109V-expressing embryos show a rightward shift, indicating increased frequency of larger transverse movement. Statistical significance was assessed using the Kolmogorov–Smirnov test on pooled data from 3 ANLN+WT embryos (n = 29 junctions) and from 3 ANLN+1109V embryos (n = 31 junctions). ***$p$<0.0001.

**Figure S10. Montage of F-actin dynamics in *X. laevis* embryos and schematic of extracellular ATP addition.**
**(A)** Time-lapse confocal imaging of F-actin (LifeAct-RFP, Fire LUT) in ANLN KD+WT or ANLN KD+1109V *X. laevis* embryos. Microscopy montage supports kymographs in Fig. 6B indicating that F-actin accumulation is often imbalanced – higher on one side than the other – for bulged junctions in ANLN KD+L1109V embryos (white arrows), while ANLN KD+WT embryos show balanced distribution of F-actin on either side of junctions throughout the time series.
**(B)** Schematic of extracellular ATP addition. Embryos are placed on a slide with an offset coverslip to allow for the addition of ATP solution during live imaging. ATP induces an increase in cortical actin contraction.

<u>**Movie Legends:**</u>

**Movie S1:** Timelapse microscopy showing F-actin (Fire LUT) in ANLN KD+WT (left) and ANLN KD+1109V (right) *X. laevis* embryos. The ANLN KD+WT embryo shows relatively uniform F-actin intensity on either side of stable, linear cell-cell junctions, while the ANLN KD+1109V embryo shows repeated, transient, imbalanced accumulations of F-actin on one side of a bulged cell-cell junction. Time interval is 22 seconds, frame rate is 7.0 fps. Related to Fig. 6.

**Movie S2:** Timelapse microscopy of ANLN KD+WT (left) and ANLN KD+1109V (right) *X. laevis* embryos expressing a probe for F-actin (LifeAct-RFP, Fire LUT) where exogenous ATP is added to the slide after two minutes of imaging (yellow box) to induce mechanical challenge. ANLN KD+1109V embryos (right panel) show an increase in transient 'flares' of F-actin following ATP treatment, Time interval is 22 seconds, frame rate is 12.0 fps. Related to Fig. 6.

**Movie S3:** Timelapse microscopy of *X. laevis* embryos during the ZnUMBA assay. ANLN KD+WT embryos (left panel) show relatively low and stable levels of junctional FluoZin3 intensity (Fire LUT) compared to ANLN KD+1109V embryos (right panel), which show increased junctional FluoZin3 intensity over time as well as local FluoZin3 increases. Time interval is 30 seconds, frame rate is 12.0 fps. Related to Fig. 7.

**Table S1.** List of known monogenic contributors to proteinuric kidney disease.

| Gene Symbol | Gene Product | Mode of Inheritance | Reference |
|---|---|---|---|
| ACTN4 | Alpha-actinin-4 | AD | PMID: 10700177 |
| ADCK4 | COQ8B, AarF domain containing kinase 4 | AR | PMID: 24270420 |
| ALG1 | Chitobiosyldiphosphodolichol beta-mannosyltransferase | AR | PMID: 27325525 |
| ANLN | Actin-binding protein anillin | AD | PMID: 24676636 |
| APOL1 | Apolipoprotein L1 | AR | PMID: 24206458 |
| ARHGAP24 | Rho GTPase-activating protein 24 | AD | PMID: 21911940 |
| ARHGDIA | Rho GDP-dissociation inhibitor 1 | AR | PMID: 23867502 |
| AVIL | Advillin | AR | PMID: 29058690 |
| CD151 | CD151 antigen | AR | PMID: 35278129 |
| CD2AP | CD2-associated protein | AR | PMID: 12764198 |
| CFH | Complement factor H | AD | PMID: 14978182 |
| CLCN5 | H(+)/Cl(-) exchange transporter 5 | X | PMID: 8559248 |
| COL4A3 | Collagen alpha-3(IV) chain | AR | PMID: 7987301 |
| COL4A4 | Collagen alpha-4(IV) chain | AR | PMID: 7987396 |
| COL4A5 | Collagen alpha-5(IV) chain | X | PMID: 8132760 |
| COQ2 | Coenzyme Q2 4-hydroxybenzoate polyprenyltransferase | AR | PMID: 17855635 |
| COQ6 | Coenzyme Q6 ubiquinone biosynthesis monooxygenase | AR | PMID: 21540551 |
| CRB2 | Protein crumbs homolog 2 | AR | PMID: 25557779 |
| CUBN | Cubilin | AR | PMID: 21903995 |
| DGKE | Diacylglycerol kinase epsilon | AR | PMID: 23274426 |
| E2F3 | Transcription factor E2F3 | AD | PMID: 21372519 |
| EMP2 | Epithelial membrane protein 2 | AR | PMID: 24814193 |
| FAT1 | Protocadherin Fat 1 | AR | PMID: 26905694 |
| HNF1B | Hepatocyte nuclear factor 1-beta | AD | PMID: 10484768 |
| INF2 | Inverted formin-2 | AD | PMID: 20023659 |
| ITGA3 | Integrin alpha-3 | AR | PMID: 25810266 |
| ITGB4 | Integrin beta-4 | AR | PMID: 10873890 |
| KANK1 | KN motif and ankyrin repeat domain-containing protein 1 | AR | PMID: 25961457 |
| KANK2 | KN motif and ankyrin repeat domain-containing protein 2 | AR | PMID: 25961457 |
| KANK4 | KN motif and ankyrin repeat domain-containing protein 4 | AR | PMID: 25961457 |
| LAGE3 | EKC/KEOPS complex subunit LAGE3 | AR | PMID: 28805828 |
| LAMB2 | Laminin subunit beta-2 | AR | PMID: 15367484 |
| LMNA | Prelamin-A/C | AD | PMID: 24080738 |

| | | | |
|---|---|---|---|
| LMX1B | LIM homeobox transcription factor 1-beta | AD | PMID: 9590287 |
| MT-TL1 | Mitochondrially encoded tRNA leucine 1 | AR | PMID: 11044204 |
| MYH9 | Myosin heavy chain 9, nonmuscle | AD | PMID: 11590545 |
| MYO1E | Unconventional myosin Ie | AR | PMID: 21756023 |
| NPHS1 | Nephrin | AR | PMID: 9660941 |
| NPHS2 | Podocin | AR | PMID: 10742096 |
| NUP93 | Nucleoporin, 93 kDa | AR | PMID: 26878725 |
| NUP107 | Nucleoporin, 107 kDa | AR | PMID: 26411495 |
| NUP205 | Nucleoporin, 205 kDa | AR | PMID: 26878725 |
| NXF5 | Nuclear RNA export factor 5 | X | PMID: 23686279 |
| OCRL | Inositol polyphosphate 5-phosphatase OCRL | X | PMID: 31811534 |
| OSGEP | tRNA N6-adenosine threonylcarbamoyltransferase | AR | PMID: 28805828 |
| PAX2 | Paired box protein Pax-2 | AD | PMID: 24676634 |
| PDSS2 | All trans-polyprenyl-diphosphate synthase PDSS2 | AR | PMID: 17186472 |
| PLCE1 | Phospholipase C, epsilon 1 | AR | PMID: 17086182 |
| PMM2 | Phosphomannomutase 2 | AR | PMID: 34546508 |
| PODXL | Podocalyxin | AD | PMID: 24048372 |
| PTPRO | Receptor-type tyrosine-protein phosphatase O | AR | PMID: 21722858 |
| SCARB2 | Lysosome membrane protein 2 | AR | PMID: 18308289 |
| SGPL1 | Sphingosine-1-phosphate lyase 1 | AR | PMID: 28165339 |
| SMARCAL1 | SWI/SNF-related matrix-associated actin-dependent regulator of chromatin subfamily A-like protein 1 | AR | PMID: 11799392 |
| SYNPO | Synaptopodin | AD | PMID: 28117080 |
| TNS2 | Tensin-2 | AR | PMID: 29773874 |
| TP53RK | EKC/KEOPS complex subunit TP53RK | AR | PMID: 28805828 |
| TPRKB | EKC/KEOPS complex subunit TPRKB | AR | PMID: 28805828 |
| TRPC6 | Short transient receptor potential channel 6 | AD | PMID: 15879175 |
| TTC21B | Tetratricopeptide repeat protein 21B | AR | PMID: 24876116 |
| UMOD | Uromodulin | AD | PMID: 32274456 |
| WDR73 | Integrator complex assembly factor WDR73 | AR | PMID: 25466283 |
| WT1 | Wilms tumor protein | AD | PMID: 1317572 |
| XPO5 | Exportin-5 | AR | PMID: 26878725 |
| ZMPSTE24 | CAAX prenyl protease 1 homolog | AR | PMID: 17152860 |

**Table S2.** List of Reactome pathway members including: RHO GTPases Activate ROCKs (R-HSA-5627117), Tight Junction interactions (R-HSA-420029), and Nephrin Family interactions (R-HSA-373753) (*60*).

| Gene Name | UniProt ID | Reactome Pathway |
|---|---|---|
| ACTN1 | P12814 | R-HSA-373753 |
| ACTN2 | P35609 | R-HSA-373753 |
| ACTN3 | Q08043 | R-HSA-373753 |
| ACTN4 | O43707 | R-HSA-373753 |
| CASK | O14936 | R-HSA-373753 |
| CD2AP | Q9Y5K6 | R-HSA-373753 |
| CFL1 | P23528 | R-HSA-5627117 |
| CLDN1 | O95832 | R-HSA-420029 |
| CLDN10 | P78369 | R-HSA-420029 |
| CLDN11 | O75508 | R-HSA-420029 |
| CLDN12 | P56749 | R-HSA-420029 |
| CLDN14 | O95500 | R-HSA-420029 |
| CLDN15 | P56746 | R-HSA-420029 |
| CLDN16 | Q9Y5I7 | R-HSA-420029 |
| CLDN17 | P56750 | R-HSA-420029 |
| CLDN18 | P56856 | R-HSA-420029 |
| CLDN19 | Q8N6F1 | R-HSA-420029 |
| CLDN2 | P57739 | R-HSA-420029 |
| CLDN20 | P56880 | R-HSA-420029 |
| CLDN22 | Q8N7P3 | R-HSA-420029 |
| CLDN23 | Q96B33 | R-HSA-420029 |
| CLDN3 | O15551 | R-HSA-420029 |
| CLDN4 | O14493 | R-HSA-420029 |
| CLDN5 | O00501 | R-HSA-420029 |
| CLDN6 | P56747 | R-HSA-420029 |
| CLDN7 | O95471 | R-HSA-420029 |
| CLDN8 | P56748 | R-HSA-420029 |
| CLDN9 | O95484 | R-HSA-420029 |
| CRB3 | Q9BUF7 | R-HSA-420029 |
| E | P0DTC4 | R-HSA-420029 |
| F11R | Q9Y624 | R-HSA-420029 |
| FYN | P06241 | R-HSA-373753 |
| IQGAP1 | P46940 | R-HSA-373753 |
| KIRREL | Q96J84 | R-HSA-373753 |
| KIRREL2 | Q6UWL6 | R-HSA-373753 |
| KIRREL3 | Q8IZU9 | R-HSA-373753 |
| LIMK1 | P53667 | R-HSA-5627117 |

| | | |
|---|---|---|
| LIMK2 | P53671 | R-HSA-5627117 |
| MAGI2 | Q86UL8 | R-HSA-373753 |
| MPP5 | Q8N3R9 | R-HSA-420029 |
| MYH10 | P35580 | R-HSA-5627117 |
| MYH11 | P35749 | R-HSA-5627117 |
| MYH14 | Q7Z406 | R-HSA-5627117 |
| MYH9 | P35579 | R-HSA-5627117 |
| MYL12B | O14950 | R-HSA-5627117 |
| MYL6 | P60660 | R-HSA-5627117 |
| MYL9 | P24844 | R-HSA-5627117 |
| NCK1 | P16333 | R-HSA-373753 |
| NCK2 | O43639 | R-HSA-373753 |
| NPHS1 | O60500 | R-HSA-373753 |
| NPHS2 | Q9NP85 | R-HSA-373753 |
| PAK1 | Q13153 | R-HSA-5627117 |
| PARD3 | Q8TEW0 | R-HSA-420029 |
| PARD6A | Q9NPB6 | R-HSA-420029 |
| PARD6B | Q9BYG5 | R-HSA-420029 |
| PARD6G | Q9BYG4 | R-HSA-420029 |
| PATJ | Q8NI35 | R-HSA-420029 |
| PIK3CA | P42336 | R-HSA-373753 |
| PIK3CB | P42338 | R-HSA-373753 |
| PIK3R1 | P27986 | R-HSA-373753 |
| PIK3R2 | O00459 | R-HSA-373753 |
| PPP1CB | P62140 | R-HSA-5627117 |
| PPP1R12A | O14974 | R-HSA-5627117 |
| PPP1R12B | O60237 | R-HSA-5627117 |
| PRKCI | P41743 | R-HSA-420029 |
| RHOA | P61586 | R-HSA-5627117 |
| RHOB | P62745 | R-HSA-5627117 |
| RHOC | P08134 | R-HSA-5627117 |
| ROCK1 | Q13464 | R-HSA-5627117 |
| ROCK2 | O75116 | R-HSA-5627117 |
| SPTAN1 | Q13813 | R-HSA-373753 |
| SPTBN1 | Q01082 | R-HSA-373753 |
| WASL | O00401 | R-HSA-373753 |

**Members of the Nephrotic Syndrome Study Network (NEPTUNE)**
**NEPTUNE Collaborating Sites**

*Atrium Health Levine Children's Hospital, Charlotte, SC*: Susan Massengill*, Layla Lo#
*Cleveland Clinic, Cleveland, OH*: Katherine Dell*, John O'Toole*, John Sedor**, Victoria Grange#
*Children's Hospital, Denver, CO:* Bradley Dixon*, Nathan Rogers#
*Children's Hospital, Los Angeles, CA*: Rachel Lestz*, Natalie Esquivias#
*Children's Mercy Hospital, Kansas City, MO*: Tarak Srivastava*, Kelsey Markus#
*Cohen Children's Hospital, New Hyde Park, NY*: Christine Sethna*, Suzanne Vento#
*Columbia University, New York, NY:* Pietro Canetta*
*Duke University Medical Center, Durham, NC:* Opeyemi Olabisi*, Rasheed Gbadegesin**, Kimberly Cicio#
*Emory University, Atlanta, GA:* Laurence Greenbaum*, Chia-shi Wang*, Chris Fan#
*The Lundquist Institute, Torrance, CA:* Sharon Adler*, Janine LaPage#
*John H Stroger Cook County Hospital, Chicago, IL:* Amatur Amarah*
*Johns Hopkins Medicine, Baltimore, MD:* Meredith Atkinson*, Ryan Hutson#
*Mayo Clinic, Rochester, MN:* John Lieske, Marie Hogan, Fernando Fervenza
*Medical University of South Carolina, Charleston, SC:* David Selewski*, Cheryl Alston#
*Montefiore Medical Center, Bronx, NY:* Kim Reidy*, Michael Ross*, Frederick Kaskel**, Patricia Flynn#
*New York University Medical Center, New York, NY:* Laura Malaga-Dieguez*, Olga Zhdanova**, Laura Jane Pehrson#, Melanie Miranda#
*The Ohio State University College of Medicine, Columbus, OH*: Salem Almaani*, Laci Roberts#
*Riley Children's Hospital of Indiana University, Indianapolis, IN:* Myda Khalid*, Veronica Servin#
*Stanford University, Stanford, CA:* Richard Lafayette*, Elizabeth Chen#
*Temple University, Philadelphia, PA:* Iris Lee**
*Texas Children's Hospital at Baylor College of Medicine, Houston, TX*: Shweta Shah*, Thinh Phan#
*University Health Network Toronto:* Heather Reich*, Michelle Hladunewich**, Paul Ling#, Martin Romano#
*University of California at San Diego, San Diego, CA:* Ambarish Athavale*, Caitlin Carter*, Kristin Zeeb#
*University of California at San Francisco, San Francisco, CA*: Paul Brakeman*, Daniel Schrader
*University of Colorado Anschutz Medical Campus, Aurora, CO*: James Dylewski* Nathan Rogers#
*University of Kansas Medical Center, Kansas City, KS*: Ellen McCarthy*, Catherine Creed#
*University of Miami, Miami, FL:* Alessia Fornoni*, Miguel Bandes#
*University of Michigan, Ann Arbor, MI:* Matthias Kretzler*, Laura Mariani*, Zubin Modi*, Amanda Williams#, Roxy Ni#
*University of Minnesota, Minneapolis, MN:* Patrick Nachman*, Michelle Rheault*, Ariel Langenberger#, Brady Wallner#
*University of North Carolina, Chapel Hill, NC:* Vimal Derebail*, Keisha Gibson*, Anne Froment#, Sharia Warren#

*University of Pennsylvania, Philadelphia, PA:* Lawrence Holzman*, Kevin Meyers**, Krishna Kallem#, Arielle Swenson#
*University of Texas San Antonio, San Antonio, TX*: Samin Sharma**
*University of Texas Southwestern, Dallas, TX:* Elizabeth Roehm*, Kamalanathan Sambandam**, Elizabeth Brown**
*University of Washington, Seattle, WA:* Ashley Jefferson*, Sangeeta Hingorani**, Katherine Tuttle**§, Linda Manahan #, Emily Pao#, Kelli Kuykendall§
*Wake Forest University Baptist Health, Winston-Salem, NC:* Jen Jar Lin**
*Washington University in St. Louis, St. Louis, MO*: Brian Stotter*, Joseph Dumayas#
**Data Analysis and Coordinating Center:** *University of Michigan:* Matthias Kretzler*, Brenda Gillespie**, Laura Mariani**, Zubin Modi**, Eloise Salmon**, Howard Trachtman**, Hailey Desmond, Sean Eddy, Damian Fermin, Wenjun Ju, Maria Larkina, Chrysta Lienczewski, Rebecca Scherr, Jonathan Troost, Amanda Williams, Yan Zhai; *Cleveland Clinic:* Crystal Gadegbeku**, John Sedor**, *Duke University:* Laura Barisoni**; *Harvard University:* Matthew G Sampson**; *Northwestern University:* Abigail Smith**; *University of Pennsylvania:* Lawrence Holzman**, Jarcy Zee**
**Digital Pathology Committee:** Carmen Avila-Casado *(University Health Network)*, Serena Bagnasco *(Johns Hopkins University)*, Lihong Bu *(Mayo Clinic)*, Shelley Caltharp *(Emory University)*, Clarissa Cassol *(Arkana)*, Dawit Demeke *(University of Michigan)*, Brenda Gillespie *(University of Michigan)*, Jared Hassler *(Temple University)*, Leal Herlitz *(Cleveland Clinic)*, Stephen Hewitt *(National Cancer Institute)*, Jeff Hodgin *(University of Michigan)*, Danni Holanda *(Arkana)*, Neeraja Kambham *(Stanford University)*, Kevin Lemley, Laura Mariani *(University of Michigan)*, Nidia Messias *(Washington University)*, Alexei Mikhailov *(Wake Forest)*, Vanessa Moreno *(University of North Carolina)*, Behzad Najafian *(University of Washington)*, Matthew Palmer *(University of Pennsylvania)*, Avi Rosenberg *(Johns Hopkins University)*, Virginie Royal *(University of Montreal)*, Miroslav Sekulik *(Columbia University)*, Barry Stokes *(Columbia University)*, David Thomas *(Duke University)*, Ming Wu *(University of New York)*, Michifumi Yamashita *(Cedar Sinai)*, Hong Yin *(Emory University)*, Jarcy Zee *(University of Pennsylvania)*, Yiqin Zuo *(University of Miami)*. Co-Chairs: Laura Barisoni *(Duke University)*, Cynthia Nast *(Cedar Sinai)*.

Fig. S1

**A**

| dbSNP ID | hg38 coord | # NEPTUNE individuals affected | gnomAD max allele frequency | DNA base change | Amino acid change | Predicted impact | Diagnosis |
|---|---|---|---|---|---|---|---|
| rs368071868 | 7:36389966 | 1 | 6.27E-04 | c.-61C>A | N/A | HIGH | Unknown |
| rs200742993 | 7:36425997 | 4 | 5.48E-03 | c.2749-4_2749 -3dup (C>CCT) | N/A | HIGH | FSGS (1); Other (2); Unknown (1) |

**B** Additional variants in individuals with ANLN VUS I1109V

| Patient | Gene | Variant | Genotype | Amino acid change | Variant Effect Predictor (VEP) | Clinical Significance (ClinVar) | gnomAD allele frequency | Category |
|---|---|---|---|---|---|---|---|---|
| 1 | NUP107 | chr12:68700850_A>G | HET | Tyr226Cys | Missense | N/A | 4.59E-05 | Monogenic |
| 1 | COQ6 | chr14:73950334_T>A | HET | p.Met1? | LOF mutation | N/A | N/A | Monogenic |
| 1 | MYH14 | chr19:50259202_A>G | HET | Glu764Gly | Missense | N/A | 4.04E-05 | Reactome |
| 2 | ACTN2 | chr1:236754025 | HET | If C-T; Arg640Cys | Missense | Uncertain significance | 1.42E-05 | Reactome |
| 3 | COL4A3 | chr2:227283820_G>A | HET | Gly904Arg | Missense | N/A | 1.24E-06 | Monogenic |
| 4 | N/A | | | | | | | |
| 5 | EMP2 | chr16:10532943_T>C | HET | Ser156Gly | Missense | N/A | 6.24E-07 | Monogenic |
| 5 | APOL1 | G1 (chr22:36265860_A>G chr22:36265988_T>G) | HET | Ser342Gly Ile384Met | Missense Missense | Conflicting interpretations of pathogenicity; risk factor | 1.14E-02 1.12E-02 | Monogenic Monogenic |
| 6 | COL4A3 | chr2:227308959_A>C | HET | Asn1508Thr | Missense | N/A | 6.20E-07 | Monogenic |
| 6 | MYH9 | chr22:36285281_T>C | HET | Lys1775Glu | Missense | Likely benign | 1.96E-03 | Monogenic |
| 7 | N/A | | | | | | | |

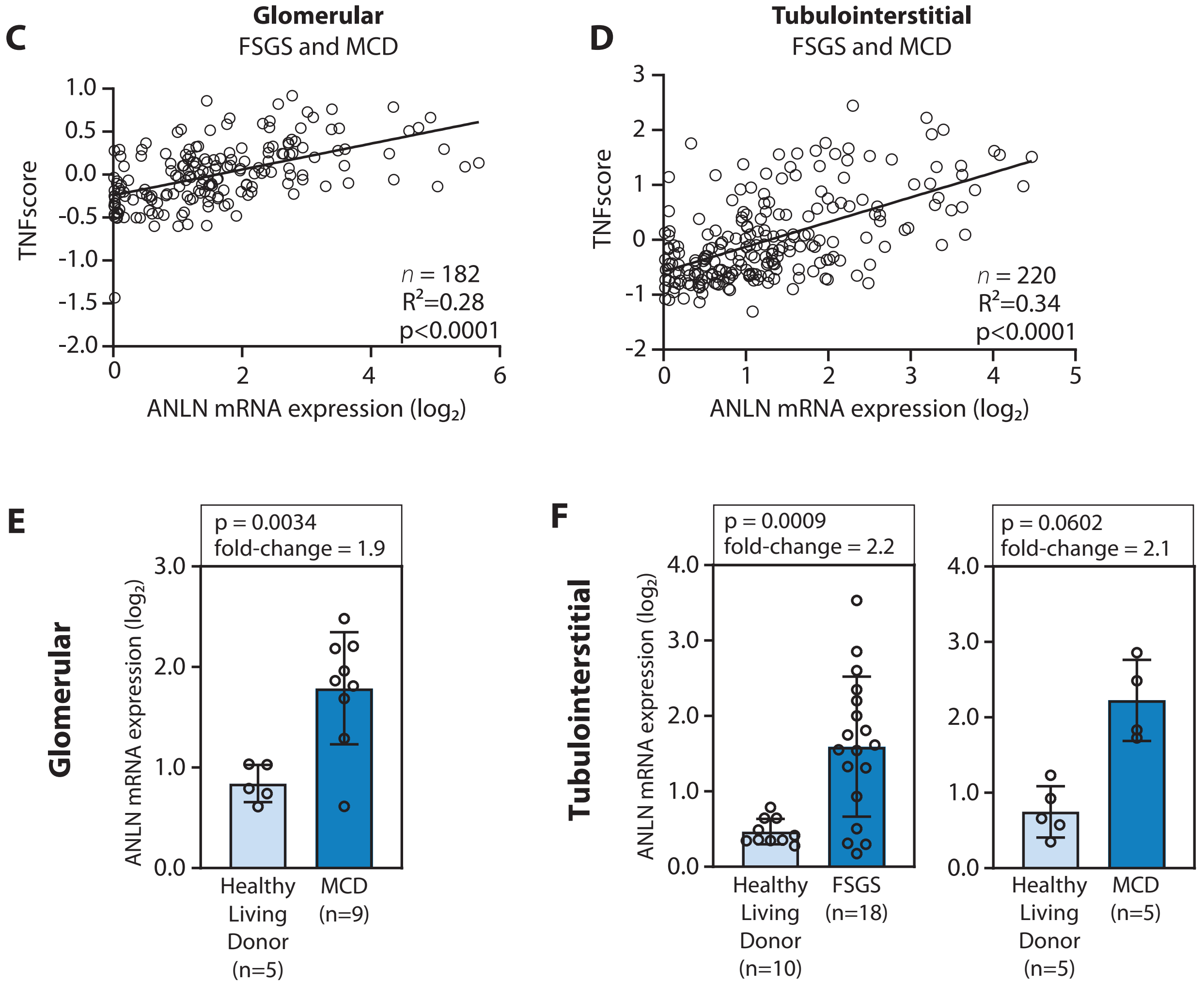

Fig. S2

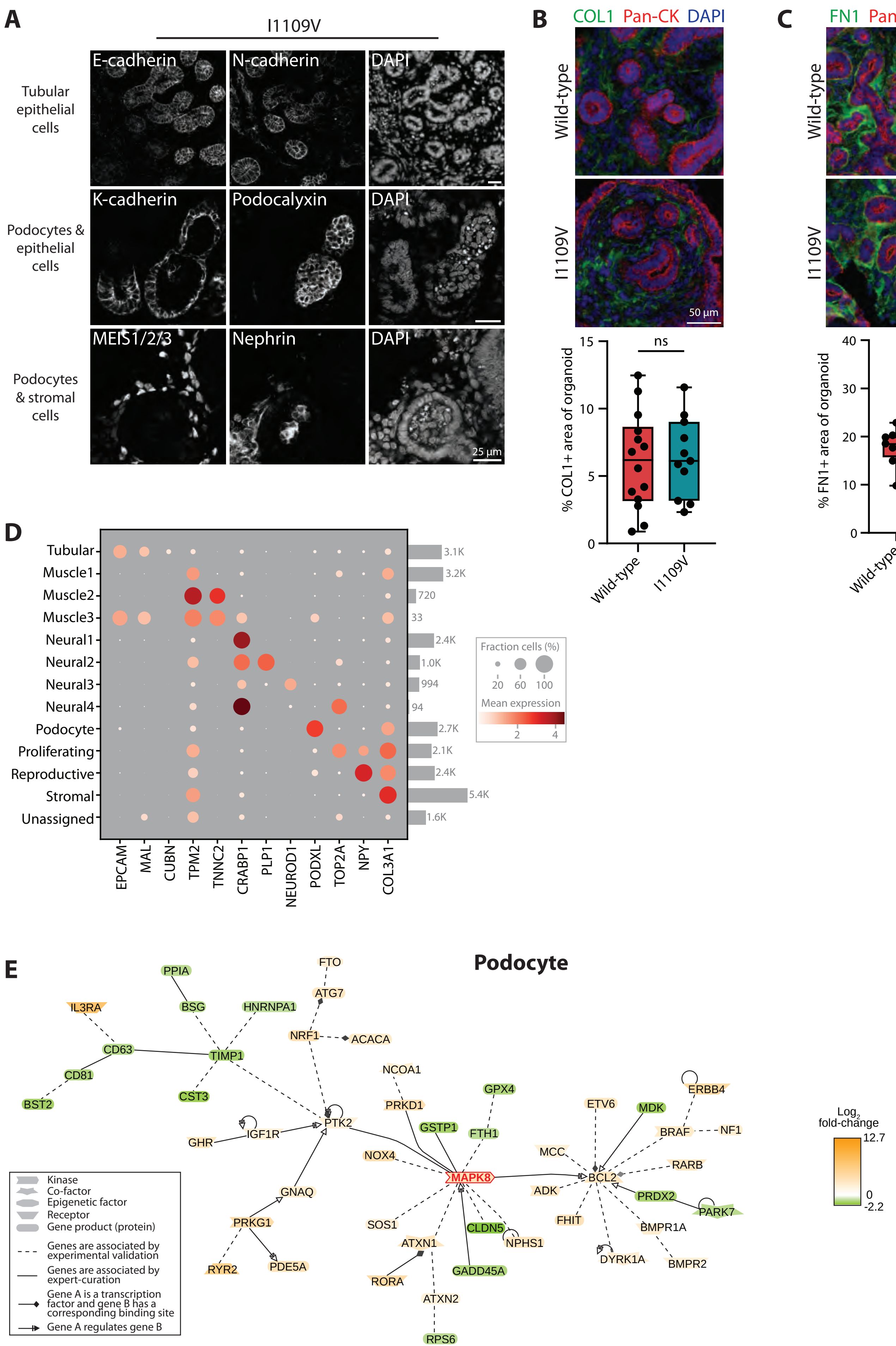
A
I1109V
Tubular epithelial cells
Podocytes & epithelial cells
Podocytes & stromal cells
E-cadherin
N-cadherin
DAPI
K-cadherin
Podocalyxin
MEIS1/2/3
Nephrin
25 µm
B
COL1 Pan-CK DAPI
Wild-type
I1109V
50 µm
% COL1+ area of organoid
ns
C
FN1 Pan-CK DAPI
% FN1+ area of organoid
D
Tubular
Muscle1
Muscle2
Muscle3
Neural1
Neural2
Neural3
Neural4
Podocyte
Proliferating
Reproductive
Stromal
Unassigned
EPCAM
MAL
CUBN
TPM2
TNNC2
CRABP1
PLP1
NEUROD1
PODXL
TOP2A
NPY
COL3A1
3.1K
3.2K
720
33
2.4K
1.0K
994
94
2.7K
2.1K
2.4K
5.4K
1.6K
Fraction cells (%)
20 60 100
Mean expression
2 4
E
Podocyte
Kinase
Co-factor
Epigenetic factor
Receptor
Gene product (protein)
Genes are associated by experimental validation
Genes are associated by expert-curation
Gene A is a transcription factor and gene B has a corresponding binding site
Gene A regulates gene B
Log2 fold-change
12.7
0
-2.2
PPIA
FTO
ATG7
IL3RA
BSG
HNRNPA1
NRF1
ACACA
CD63
TIMP1
CD81
CST3
BST2
NCOA1
PRKD1
GPX4
PTK2
GHR
IGF1R
GSTP1
FTH1
NOX4
MAPK8
GNAQ
PRKG1
SOS1
ATXN1
CLDN5
NPHS1
RYR2
PDE5A
RORA
GADD45A
ATXN2
RPS6
ETV6
MDK
ERBB4
BRAF
NF1
MCC
RARB
BCL2
ADK
PRDX2
PARK7
FHIT
BMPR1A
DYRK1A
BMPR2

Fig. S3

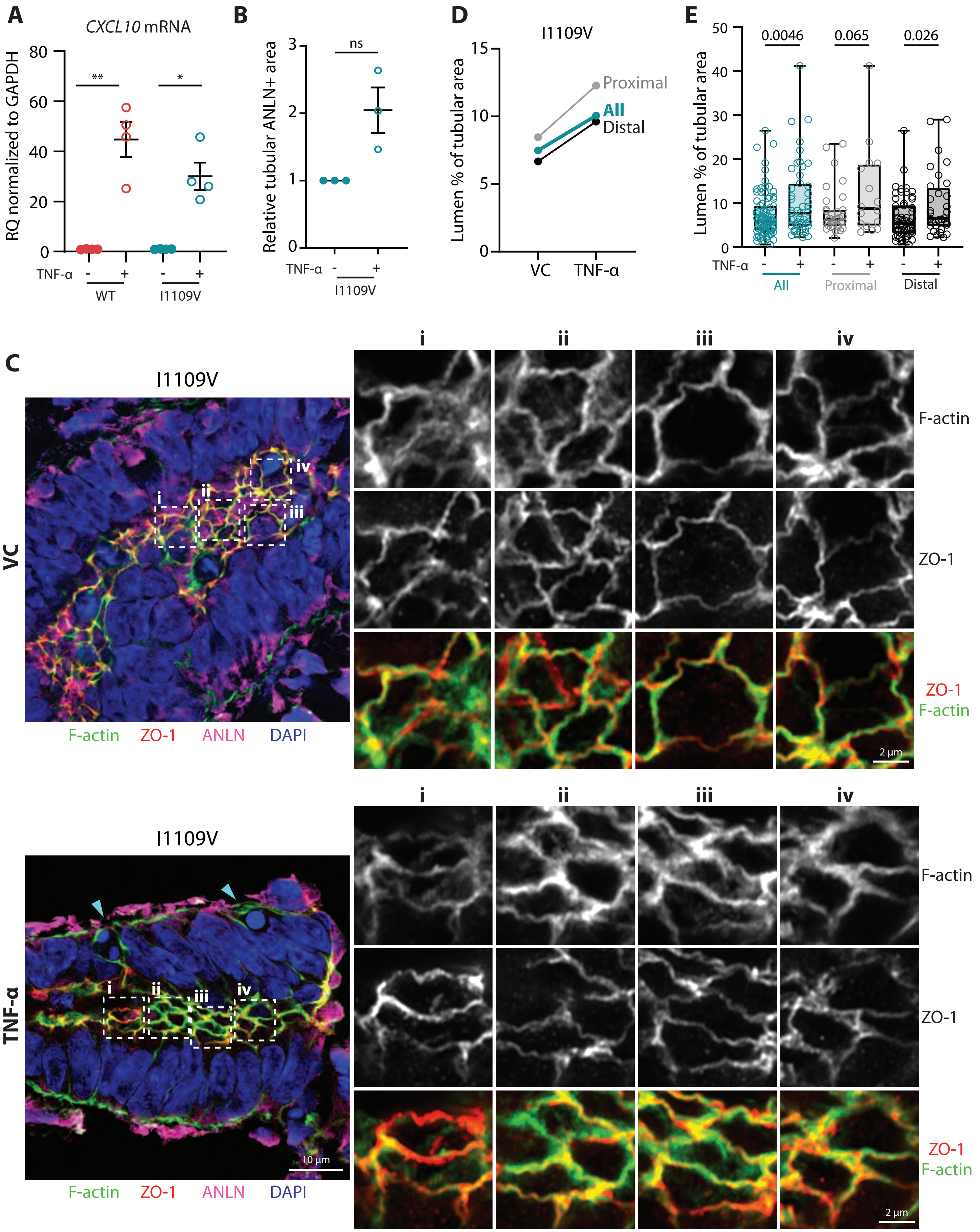
A
CXCL10 mRNA
RQ normalized to GAPDH
80
60
40
20
0
**
*
TNF-α
-
+
-
+
WT
I1109V
B
Relative tubular ANLN+ area
3
2
1
0
ns
TNF-α
-
+
I1109V
D
I1109V
Lumen % of tubular area
15
10
5
0
Proximal
All
Distal
VC
TNF-α
E
Lumen % of tubular area
50
40
30
20
10
0
0.0046
0.065
0.026
TNF-α
-
+
-
+
-
+
All
Proximal
Distal
C
I1109V
VC
i
ii
iii
iv
F-actin
ZO-1
ANLN
DAPI
F-actin
ZO-1
ZO-1
F-actin
2 µm
I1109V
TNF-α
i
ii
iii
iv
10 µm
F-actin
ZO-1
ANLN
DAPI
F-actin
ZO-1
ZO-1
F-actin
2 µm

Fig. S4

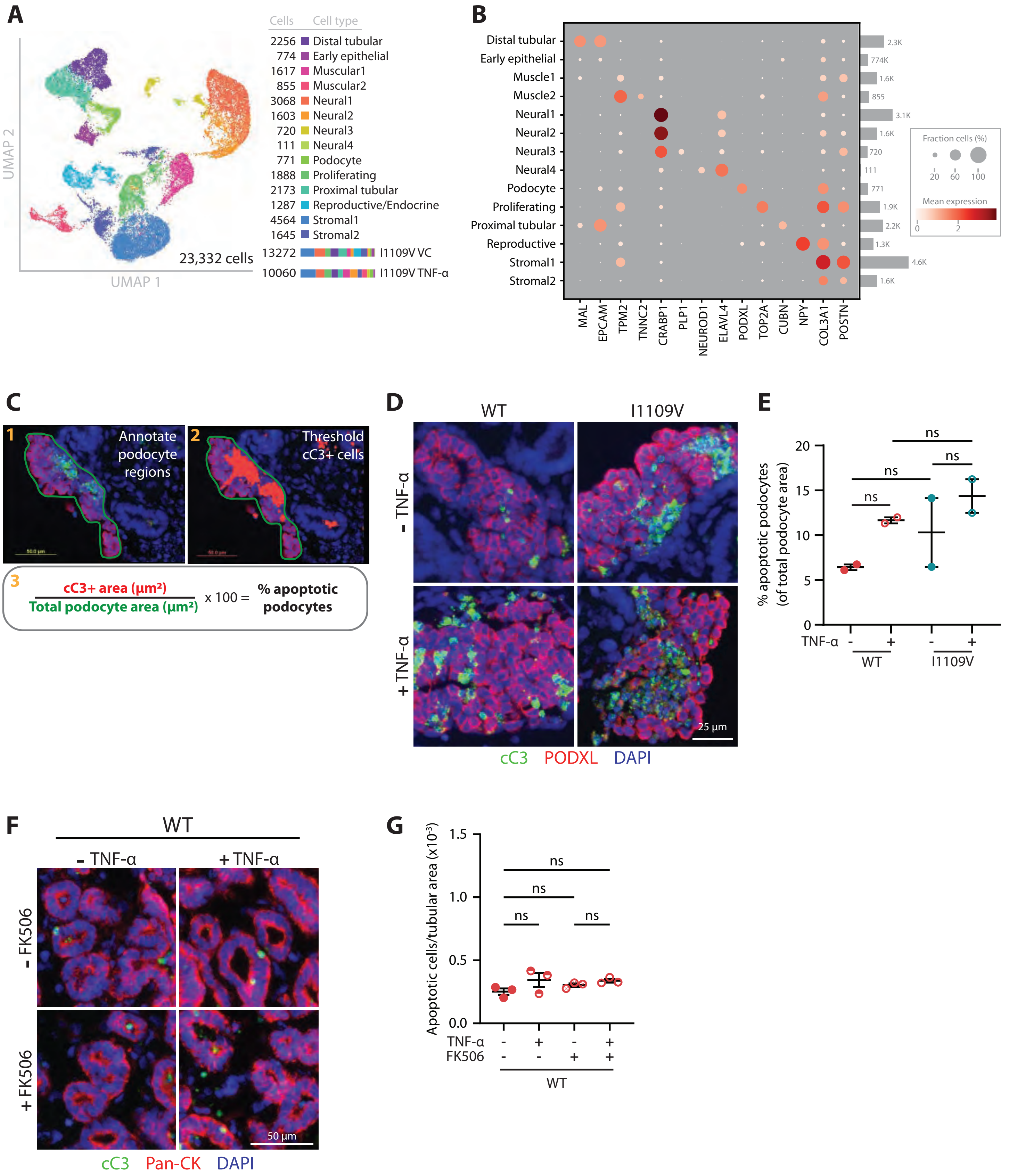
A
Cells Cell type
2256 Distal tubular
774 Early epithelial
1617 Muscular1
855 Muscular2
3068 Neural1
1603 Neural2
720 Neural3
111 Neural4
771 Podocyte
1888 Proliferating
2173 Proximal tubular
1287 Reproductive/Endocrine
4564 Stromal1
1645 Stromal2
13272 I1109V VC
10060 I1109V TNF-α
23,332 cells
UMAP 1
UMAP 2
B
Distal tubular
Early epithelial
Muscle1
Muscle2
Neural1
Neural2
Neural3
Neural4
Podocyte
Proliferating
Proximal tubular
Reproductive
Stromal1
Stromal2
MAL
EPCAM
TPM2
TNNC2
CRABP1
PLP1
NEUROD1
ELAVL4
PODXL
TOP2A
CUBN
NPY
COL3A1
POSTN
Fraction cells (%)
20 60 100
Mean expression
0 2
C
1 Annotate podocyte regions
2 Threshold cC3+ cells
3 cC3+ area (µm²) / Total podocyte area (µm²) x 100 = % apoptotic podocytes
D
WT
I1109V
- TNF-α
+ TNF-α
25 µm
cC3 PODXL DAPI
E
% apoptotic podocytes (of total podocyte area)
ns
TNF-α - + - +
WT
I1109V
F
WT
- TNF-α
+ TNF-α
- FK506
+ FK506
50 µm
cC3 Pan-CK DAPI
G
Apoptotic cells/tubular area (x10⁻³)
ns
TNF-α - + - +
FK506 - - + +
WT

Fig. S5

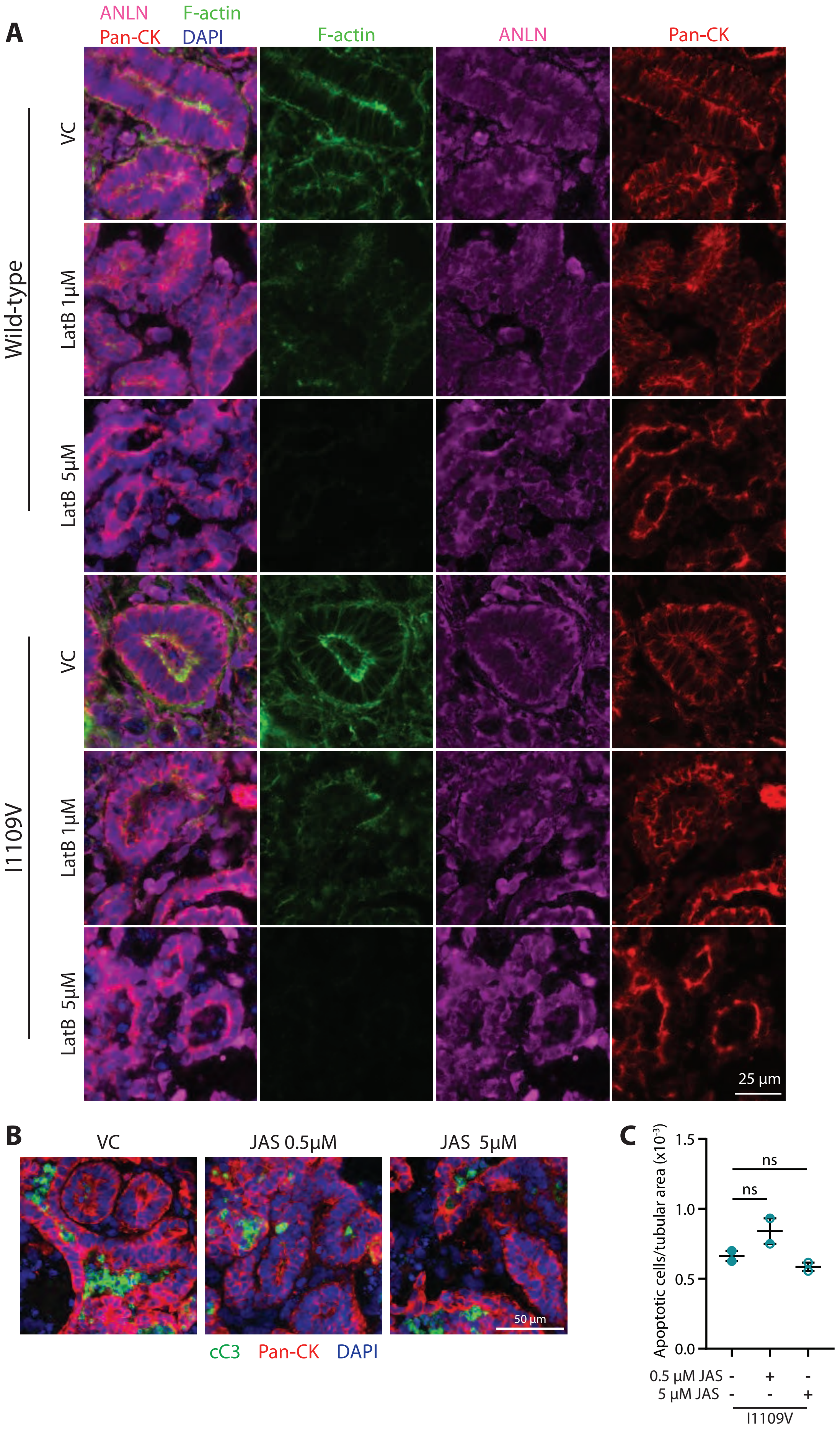
A
ANLN F-actin
Pan-CK DAPI
F-actin
ANLN
Pan-CK
Wild-type
VC
LatB 1μM
LatB 5μM
I1109V
VC
LatB 1μM
LatB 5μM
25 μm
B
VC
JAS 0.5μM
JAS 5μM
50 μm
cC3 Pan-CK DAPI
C
Apoptotic cells/tubular area (x10⁻³)
1.5
1.0
0.5
0.0
ns
ns
0.5 μM JAS - + -
5 μM JAS - - +
I1109V

Fig. S6

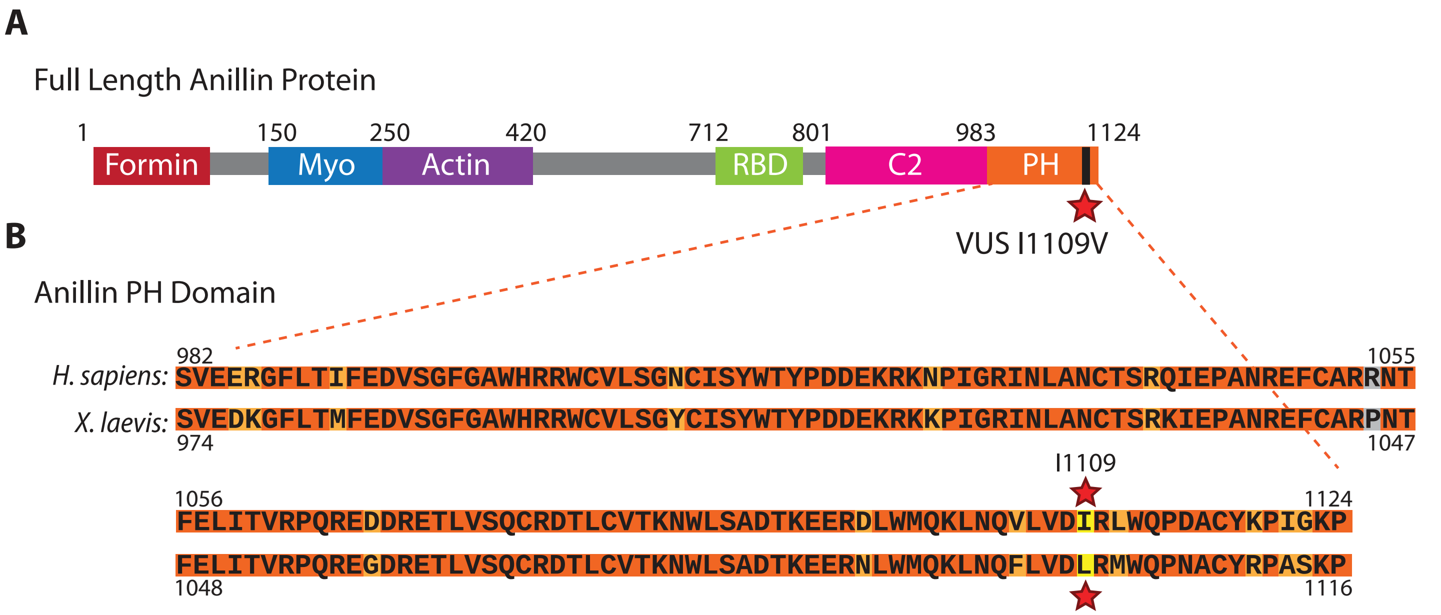


**C** Human vs. *Xenopus laevis*

| | Full Length Protein | PH Domain |
|---|---|---|
| Identity | 57.80% | 88.6% |
| Similarity | 70.9% | 95.0% |

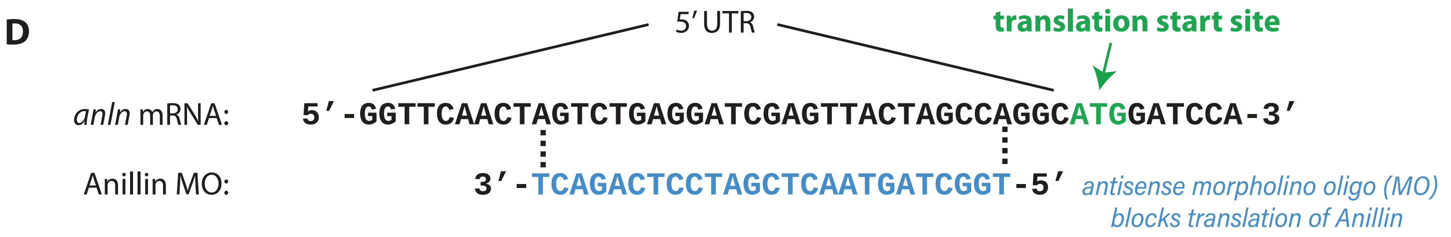


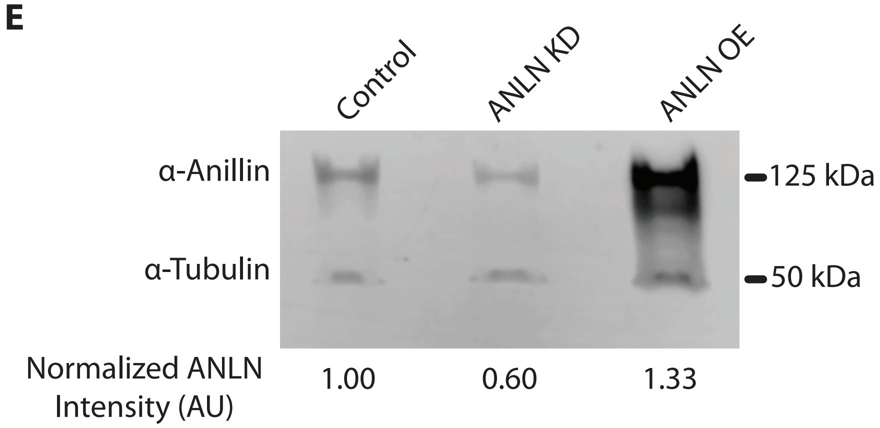

Fig. S7

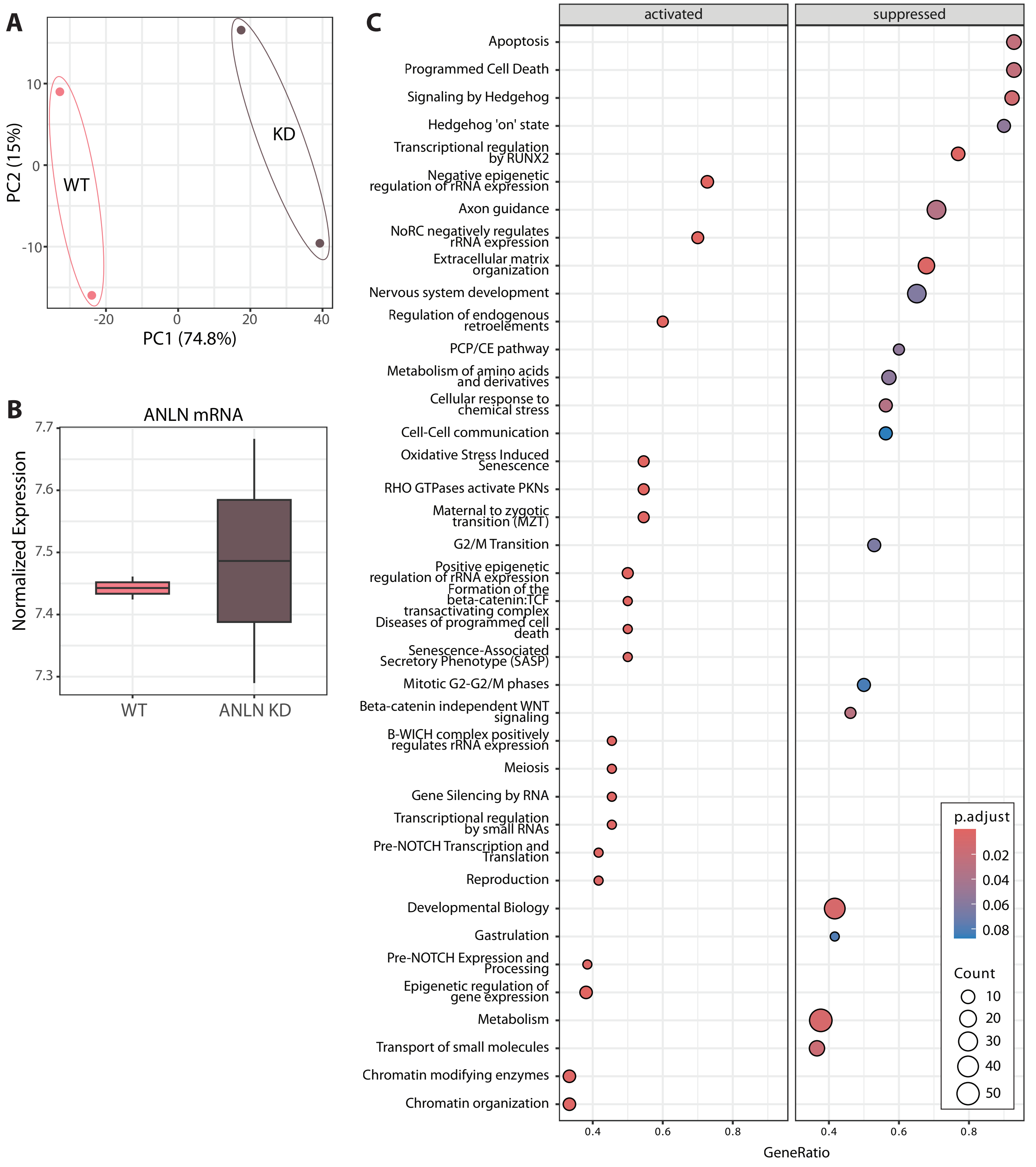

A
PC2 (15%)
PC1 (74.8%)
WT
KD
B
ANLN mRNA
Normalized Expression
WT
ANLN KD
C
activated
suppressed
Apoptosis
Programmed Cell Death
Signaling by Hedgehog
Hedgehog 'on' state
Transcriptional regulation by RUNX2
Negative epigenetic regulation of rRNA expression
Axon guidance
NoRC negatively regulates rRNA expression
Extracellular matrix organization
Nervous system development
Regulation of endogenous retroelements
PCP/CE pathway
Metabolism of amino acids and derivatives
Cellular response to chemical stress
Cell-Cell communication
Oxidative Stress Induced Senescence
RHO GTPases activate PKNs
Maternal to zygotic transition (MZT)
G2/M Transition
Positive epigenetic regulation of rRNA expression
Formation of the beta-catenin:TCF transactivating complex
Diseases of programmed cell death
Senescence-Associated Secretory Phenotype (SASP)
Mitotic G2-G2/M phases
Beta-catenin independent WNT signaling
B-WICH complex positively regulates rRNA expression
Meiosis
Gene Silencing by RNA
Transcriptional regulation by small RNAs
Pre-NOTCH Transcription and Translation
Reproduction
Developmental Biology
Gastrulation
Pre-NOTCH Expression and Processing
Epigenetic regulation of gene expression
Metabolism
Transport of small molecules
Chromatin modifying enzymes
Chromatin organization
GeneRatio
p.adjust
Count

Fig. S8

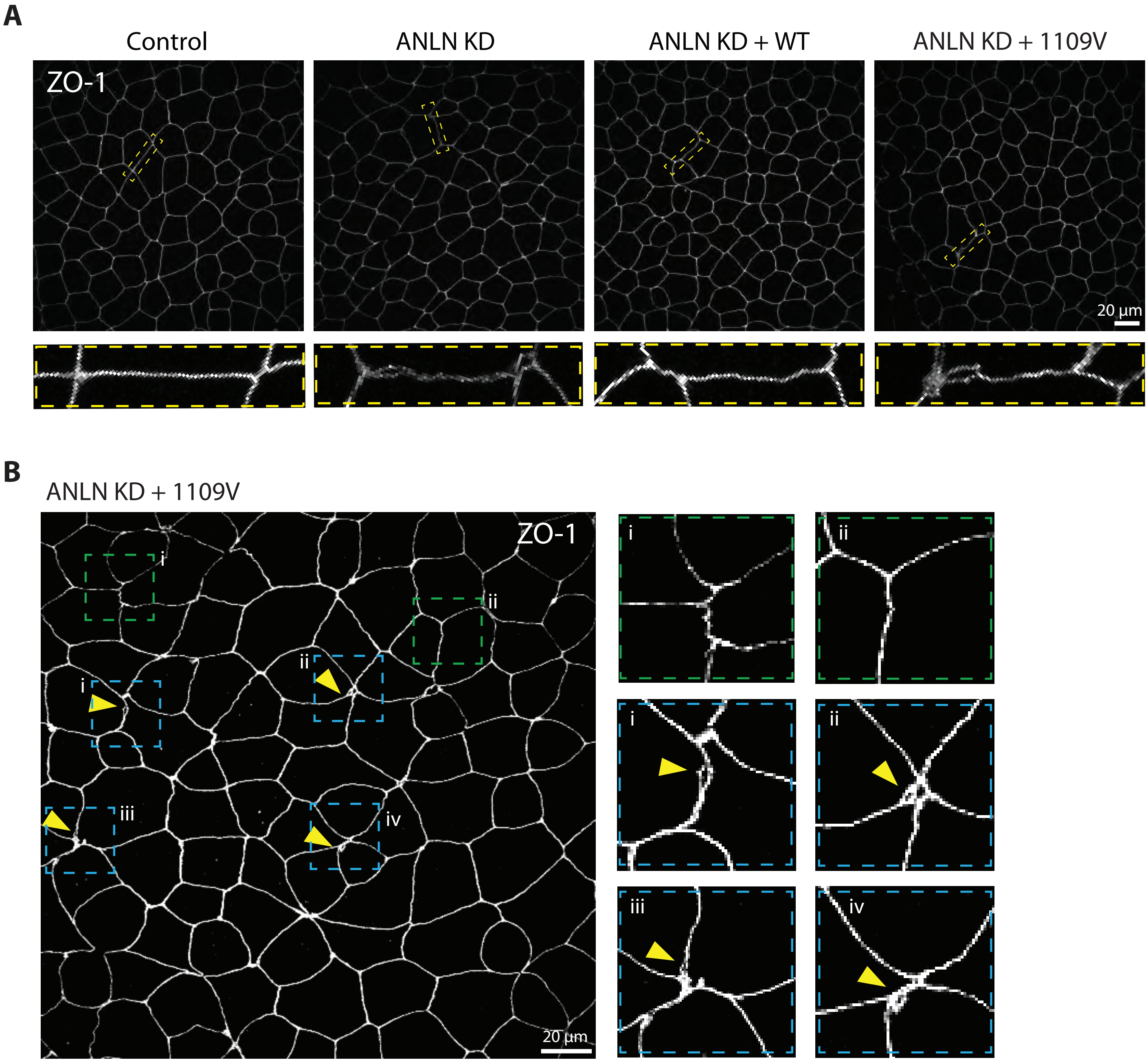
A
Control
ANLN KD
ANLN KD + WT
ANLN KD + 1109V
ZO-1
20 µm
B
ANLN KD + 1109V
ZO-1
i
ii
i
ii
iii
iv
20 µm
i
ii
i
ii
iii
iv

Fig. S9

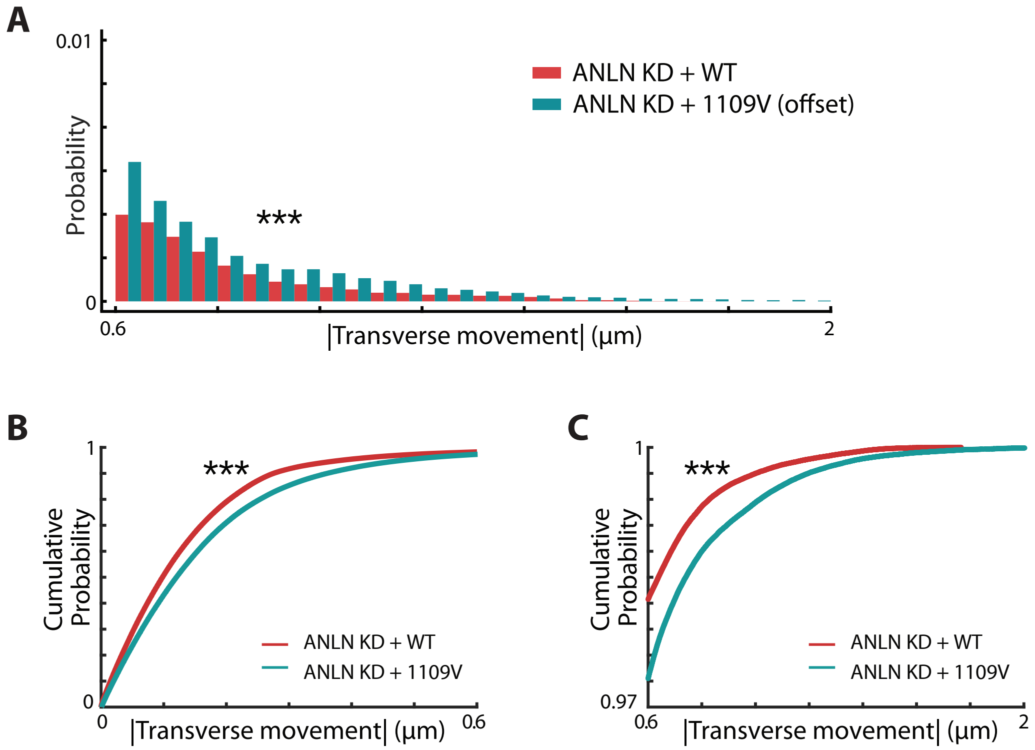
A
0.01
Probability
0
0.6
|Transverse movement| (μm)
2
***
ANLN KD + WT
ANLN KD + 1109V (offset)
B
1
Cumulative Probability
0
0
|Transverse movement| (μm)
0.6
***
ANLN KD + WT
ANLN KD + 1109V
C
1
Cumulative Probability
0.97
0.6
|Transverse movement| (μm)
2
***
ANLN KD + WT
ANLN KD + 1109V

Fig. S10

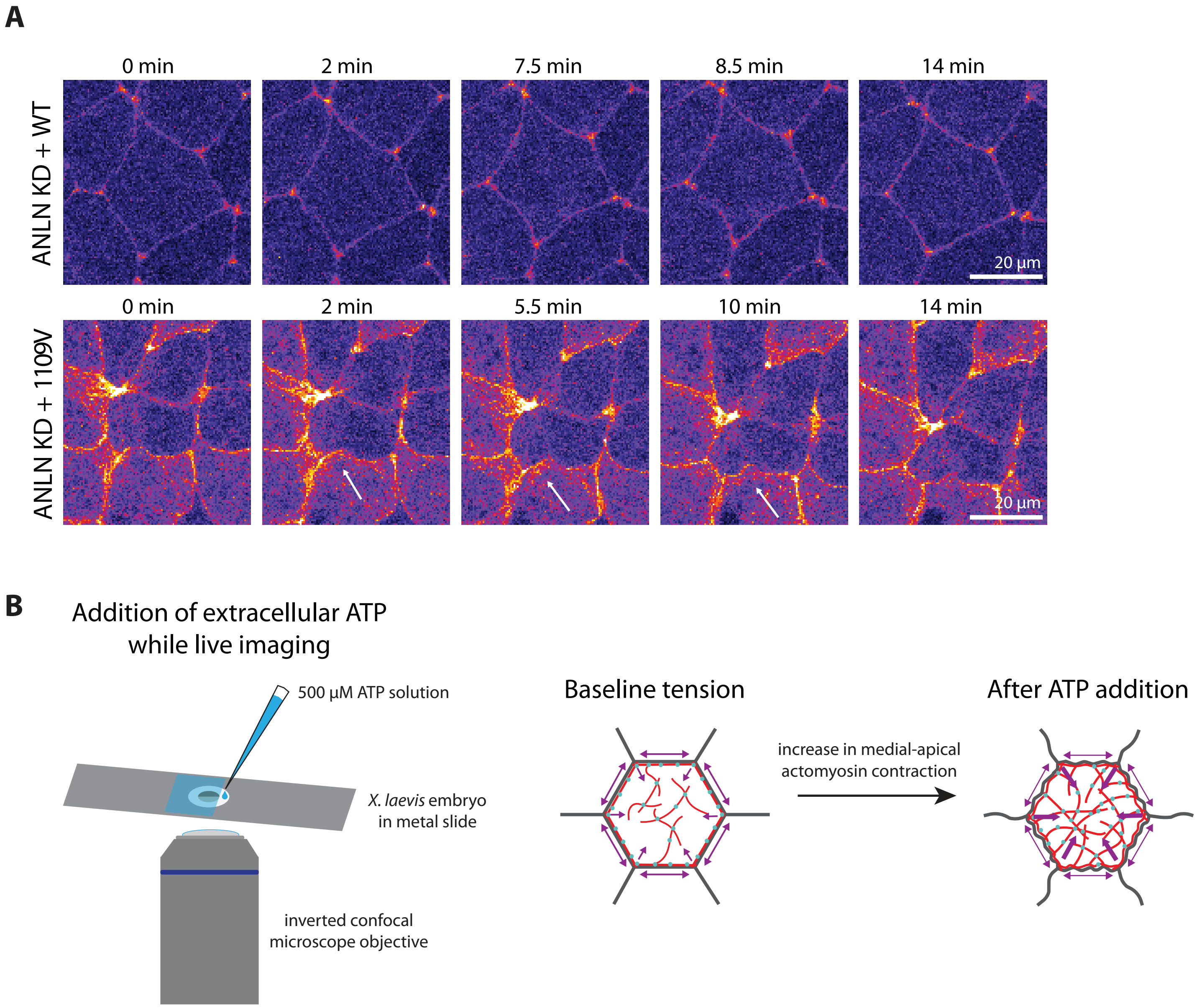
A
0 min
2 min
7.5 min
8.5 min
14 min
ANLN KD + WT
20 µm
0 min
2 min
5.5 min
10 min
14 min
ANLN KD + 1109V
20 µm
B
Addition of extracellular ATP while live imaging
500 µM ATP solution
X. laevis embryo in metal slide
inverted confocal microscope objective
Baseline tension
increase in medial-apical actomyosin contraction
After ATP addition
